# The impact of lag time to cancer diagnosis and treatment on clinical outcomes prior to the COVID-19 pandemic: a scoping review of systematic reviews and meta-analyses

**DOI:** 10.1101/2022.06.21.22276569

**Authors:** Parker Tope, Eliya Farah, Rami Ali, Mariam El-Zein, Wilson H. Miller, Eduardo L. Franco

## Abstract

**Background:** The COVID-19 pandemic has disrupted cancer care, raising concerns regarding the impact of wait time, or ‘lag time’, on clinical outcomes. We aimed to contextualize pandemic-related lag times by mapping pre-pandemic evidence from systematic reviews and/or meta-analyses on the association between lag time to cancer diagnosis and treatment with mortality- and morbidity-related outcomes.

**Methods:** We systematically searched MEDLINE, EMBASE, Web of Science, and Cochrane Library of Systematic Reviews for reviews published prior to the pandemic (1 January 2010-31 December 2019). We extracted data on methodological characteristics, lag time interval start and endpoints, qualitative findings from systematic reviews, and pooled risk estimates of mortality-(i.e., overall survival) and morbidity- (i.e., local regional control) related outcomes from meta-analyses. We categorized lag times according to milestones across the cancer care continuum and summarized outcomes by cancer site and lag time interval.

**Results:** We identified 9,032 records through database searches, of which 29 were eligible. We classified 33 unique types of lag time intervals across 10 cancer sites, of which breast, colorectal, head and neck, and ovarian cancers were investigated most. Two systematic reviews investigating lag time to diagnosis reported contradictory findings regarding survival outcomes among pediatric patients with Ewing’s sarcomas or central nervous system tumours. Comparable risk estimates of mortality were found for lag time intervals from surgery to adjuvant chemotherapy for breast, colorectal, and ovarian cancers. Risk estimates of pathologic complete response indicated an optimal time window of 7-8 weeks for neoadjuvant chemotherapy completion prior to surgery for rectal cancers. In comparing methods across meta-analyses on the same cancer sites, lag times, and outcomes, we identified critical variations in lag time research design.

**Conclusions:** Our review highlighted measured associations between lag time and cancer-related outcomes and identified the need for a standardized methodological approach in areas such as lag time definitions and accounting for the waiting-time paradox. Prioritization of lag time research is integral for revised cancer care guidelines under pandemic contingency and assessing the pandemic’s long-term effect on patients with cancer.

## INTRODUCTION

The sudden toll of the coronavirus disease of 2019 (COVID-19) pandemic on healthcare systems worldwide has transformed the provision of cancer control and care services. With successive waves of SARS-CoV-2 variants waxing and waning disparately between and within countries, the standard cancer care framework from diagnosis to treatment has been distorted. Resulting stressors on cancer centres have introduced a multitude of challenges, including prolongation of existing lag times within the cancer care continuum. In the early months of the pandemic, cancer screening services were temporarily suspended (1-3). By May 2020, two months after the World Health Organization declared the COVID-19 outbreak a pandemic, the volume of colorectal and breast cancer screenings in the United States dropped by 85% and 94%, respectively, compared to averages from the previous three years (1). Routine and diagnostic patient visits to primary healthcare physicians have been similarly affected. Clinics’ reduced overall patient volume and patient hesitancy to seek in-person care have further contributed to the backlog of elective and non-elective cancer services and procedures (4-6). New and previously diagnosed cancer cases have been subject to risk-based prioritization and treatment triaging that differ from standard clinical practice (7). For cancers that were diagnosed in the first six months of the pandemic, an increasing trend in diagnosis of late-stage cancers and a decreasing trend in diagnosis of early-stage cancers were observed, as a reflection of the initial effects of extended lag times to diagnosis (8).

Changes in dosing and fractionation, as well as delays and interruptions in chemotherapy and radiotherapy regimens of palliative and curative intent care have altered the sensitivity and timeliness of treatment administration (9, 10). In addition, COVID-19 safety measures within hospitals and cancer centers have congested surgical windows, leaving surgical treatment opportunities for only the most urgent, non-elective cases (11). Delay or cessation of clinical trials have restricted treatment and research opportunities integral for both present and future patients with cancer (12). Cumulatively, pandemic-dictated modifications to standard protocols for radiologic, surgical, and systemic therapy interventions have exacerbated lag times to cancer treatment.

Pandemic-induced changes have brought forth the rising concern within the cancer care community as to whether current lag times to diagnosis and treatment that deviate from standard-of-care practice will lead to poorer outcomes for cancer patients. With modelling studies forecasting the tolerability of these lag times based on estimated long-term outcomes (13-15) and a recent scoping review summarizing the impact of the pandemic on time to cancer diagnosis and treatment (8), a pre-pandemic perspective of the effect of lag time on oncologic outcomes among patients undergoing cancer screening, diagnosis, and staging is of paramount importance. Retrospective, pre-pandemic data can help our understanding of the influence of lag time on patient outcomes, which would be imperative for planning cancer control and care services in the future. Thus, the purpose of this scoping review is to contextualize pandemic-related lag times to cancer diagnosis and treatment by presenting an overview of aggregated pre-pandemic data from systematic reviews and/or meta-analyses on the association between lag time to cancer diagnosis and treatment and clinical outcomes.

## METHODS

Results from this review were reported in accordance with the Preferred Reporting Items for Systematic Reviews and Meta-analyses extension for Scoping Reviews (PRISMA-ScR) guidelines (16).

### Search strategy and selection criteria

We systematically searched four electronic databases: MEDLINE, EMBASE, Web of Science, and the Cochrane Library of Systematic Reviews. The search strategy consisted of the index keywords *cancer, diagnosis* & *treatment, wait-time, delay, outcome*, and *systematic review* and/or *meta-analysis*, along with their associated MeSH and iterative search terms (**Supplementary Table 1**). We queried these databases for records published between 1 January 2010 and 31 December 2019. We chose the former date to avoid capturing changes in treatment modalities for cancer sites that might have affected clinical outcomes, and the latter date to prevent artifacts from the COVID-19 pandemic era impacting our search findings. We did not apply language restrictions. In addition, we manually searched the reference lists of eligible systematic reviews and/or meta-analyses to identify potentially relevant reviews missed in our search.

### Eligibility assessment

We imported all records into EndNote X9 reference management software where duplicates were removed. Subsequently, remaining records were uploaded to Rayyan web tool for systematic reviews (17), where additional duplicates were removed. To be included, a review needed to [1] be a systematic review and/or meta-analysis, [2] refer to any clinical outcome associated with a lag time to cancer diagnosis or treatment, and [3] mention lag time to cancer diagnosis or treatment. In the first round, two reviewers (PT and EF; PT and RA; or EF and RA) independently screened the records’ titles and abstracts for eligibility in Rayyan. Discrepancies were resolved through discussion. Full-text eligibility screening was performed independently by two reviewers (EF and RA) and validated by a third (PT). Reference lists of records included in the full-text eligibility screening were manually searched by two reviewers (EF and RA) and validated by a third (PT) for further records that met the eligibility criteria.

### Data abstraction

The included records were divided into two sets and data were independently abstracted by two reviewers (PT and EF for one set, and PT and RA for the second set). One reviewer (PT) then verified all abstracted data. Inconsistencies among reviewers were resolved through discussion. From all included literature, we extracted the following variables: databases searched, number of hits, number of included studies, total number of participants, countries in which studies were conducted, start and endpoints of lag times evaluated, range of the lag time interval, cancer site, cancer type, and outcomes (e.g., mortality, disease-free survival (DFS), relapse-free survival (RFS), disease progression, etc.). We extracted additional information and variables particular to the review type. This included overall qualitative findings from systematic reviews without a meta-analytic component (referred to herein as systematic reviews). For reviews with a meta-analytic component (referred to hereafter as meta-analyses), we further extracted lag time variable type (categorical or continuous), lag time comparator and reference categories, corresponding pooled risk estimates (e.g., risk ratios (RR), hazard ratios (HR), odds ratios (OR)) with their respective 95% confidence intervals (CI), model parameters, heterogeneity statistics, and results from subgroup (by cancer type, study quality, follow-up period, and confounding adjustment, i.e., for sex, smoking, alcohol consumption, study design, and sociodemographic variables) and/or sensitivity analyses. Findings from meta-analyses were categorized by outcome: morbidity- (e.g., disease progression) or mortality-related.

### Definition and representation of lag time intervals

Based on the extracted start and endpoints of lag times, we defined unique lag time intervals that encompassed diagnostic, system, and treatment wait times. We visually represented these lag time intervals on a timeline by including horizontal bars that align the start and endpoints in relation to relevant “milestones” along the cancer care continuum (i.e., symptom onset, first seen by primary care physician, referral to a specialist, diagnosis, treatment, and palliative care). For reviews that referred to a lag time of interest using terms of common usage in the cancer community (e.g., diagnostic delay, treatment delay), we abstracted the start and endpoints of lag time intervals from the original studies and cross-validated these definitions with those used in the included review (18-20). Some reviews did not mention whether the time between the start and endpoints included or excluded other “milestones” along the cancer care continuum (20-24). We used orange shading to differentiate these from lag time intervals that included milestones in between start and endpoints which are shaded in blue (18-20, 23, 25-45).

## RESULTS

**Figure 1** outlines the search results for relevant literature and review selection. Of 9,032 records identified, 3,621 duplicates were removed, leaving 5,411 records for primary screening. Based on title and abstract screening, we excluded 5,338 records that did not meet the eligibility criteria. We retrieved the full text of 73 records and further excluded 44. We did not identify any eligible records via manual search of the reference lists of the 73 eligible records. A total of 20 meta-analyses and nine systematic reviews were included in the current scoping review.

**Figure 1.**
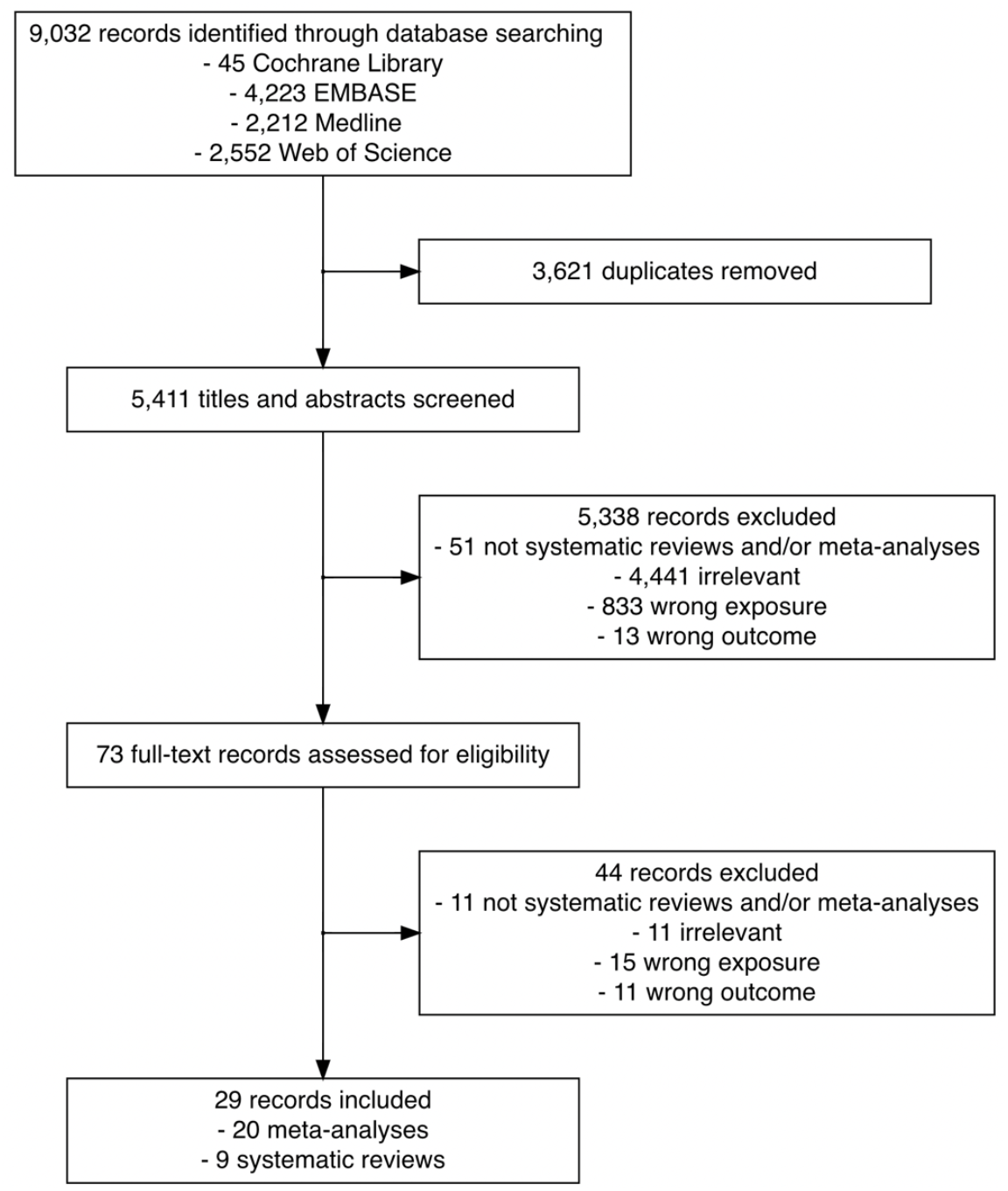
Flowchart of the search and study selection of systematic reviews and meta-analyses on the association between time to cancer diagnosis and/or treatment and clinical outcomes

As illustrated in **Figure 2**, we labelled 32 unique lag time intervals. A plurality of the identified lag time intervals encompassed lag time experienced during treatment. Notably, few reviews specified whether common milestones in care were experienced between defined start and endpoints. For example, although T18 and T22 had similar start and endpoints (from diagnosis to primary intervention by surgery, respectively), T18 is depicted in orange as it was specifically stated that studies were excluded from the review if patients underwent neoadjuvant therapy after diagnosis and before surgery (24), while T22 is depicted in blue as the corresponding review included studies of patients who underwent neoadjuvant therapy prior to surgery (20).

**Figure 2.**
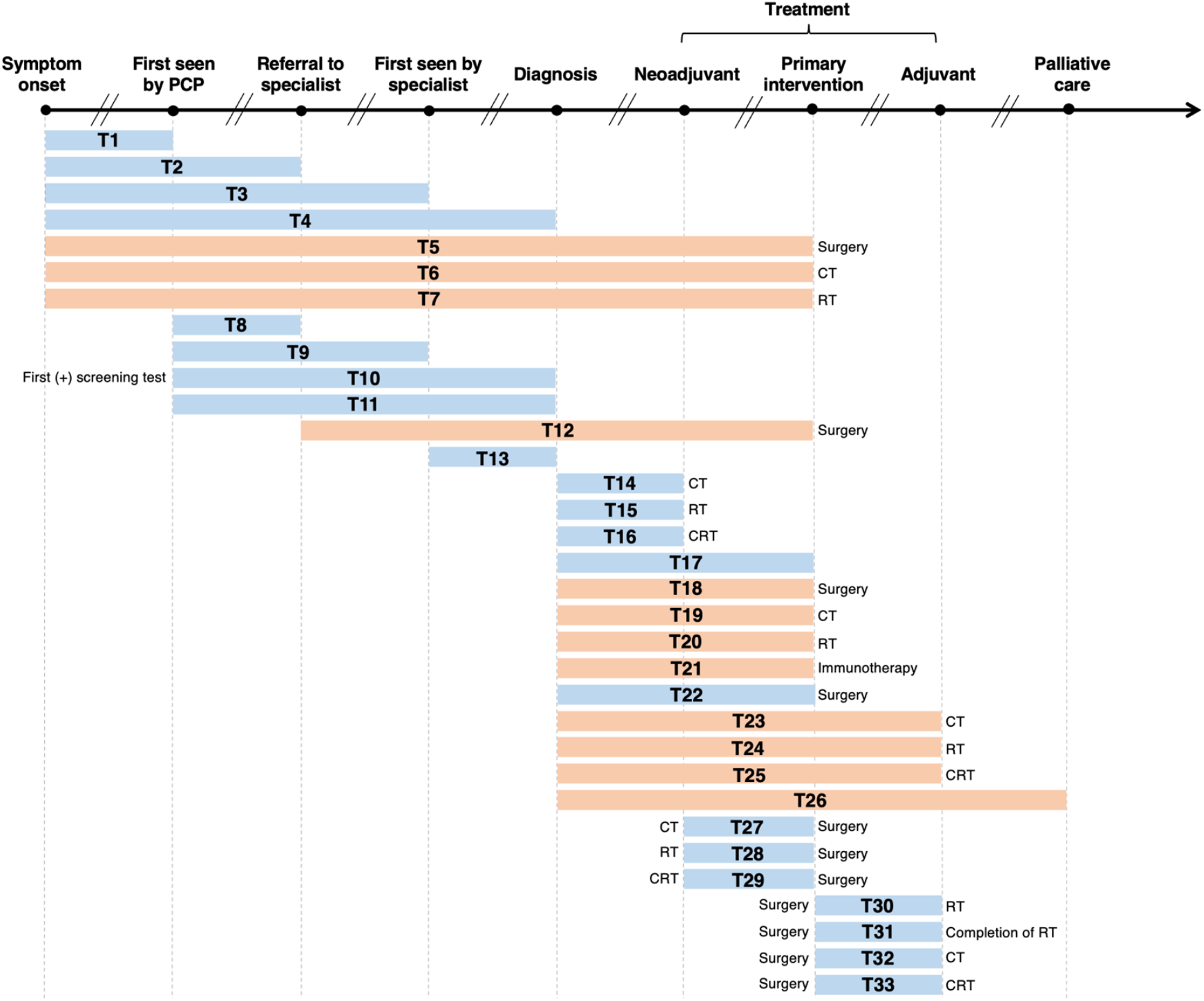
Visualization of lag time intervals identified in systematic reviews and/or meta-analyses on the association between time to cancer diagnosis and/or treatment and clinical outcomes Top arrow represents the cancer care continuum along broad milestones (data points in bold). Oblique breaks denote the incongruency of lag times between milestones (i.e., inconsistent periods of time between milestones; not every cancer patient undergoes all milestones or undergo each milestone sequentially). Each bar indicates a lag time interval (T1-T33). Start and endpoints of each lag time interval are defined by the corresponding milestones. Text before or after a bar defines specific start or endpoints of a lag time interval whenever explicitly reported in a systematic review and/or meta-analysis. Orange shading of bars denotes lag time intervals that do not necessarily include all the milestones through which the bars physically pass (e.g., T18 starts at diagnosis and ends at surgery, without necessarily including neoadjuvant therapy). Blue shading of bars denotes lag time intervals that do include all milestones through which the bars physically pass (e.g., T22 starts at diagnosis and ends at surgery, including neoadjuvant therapy). ART, adjuvant radiotherapy; CRT, chemoradiotherapy; CT, chemotherapy; PCP, primary care provider; RT, radiotherapy.

**Table 1** lists, by cancer site, the lag time intervals shown in Figure 2, where a clear pattern can be observed. Most meta-analyses on breast cancer examined the time between surgery and adjuvant chemotherapy (T32) (40-42). For colorectal cancer, the most frequently investigated lag time interval was T29 (time between neoadjuvant chemoradiotherapy and surgery) (26, 33, 34, 36). Only five records considered more than one lag time type (18, 19, 23, 24), with the broadest scope considered by Neal et al., who explored 25 unique lag time intervals across 25 cancer types (20).

**Table 1.** Lag time intervals evaluated in systematic reviews and meta-analyses on the association between time to diagnosis and/or treatment and clinical outcomes, by cancer site

| **Cancer site** | **Lag time interval<sup>a</sup>** | **First author (year), type of review** |
| --- | --- | --- |
| **Brain** | T30 | Loureiro (2016), meta-analysis (39) |
|  | T33 | Warren (2019), systematic review (25) |
| **Breast** | T32 | Yu (2013), meta-analysis (40) |
|  | T30 | Gupta (2016), meta-analysis (31) |
|  | T32 | Raphael (2016), meta-analysis (41) |
|  | T32 | Zhan (2018), meta-analysis (42) |
| **Blood** | T21 | Zhao (2019), meta-analysis (21) |
| **Colorectal** | T32 | Des Guetz (2010), meta-analysis (43) |
|  | T32 | Biagi (2011), meta-analysis (44) |
|  | T29 | Foster (2013), systematic review (26) |
|  | T29 | Wang (2015), meta-analysis (33) |
|  | T29 | Petrelli (2016), meta-analysis (34) |
|  | T29 | Du (2017), meta-analysis (36) |
|  | T28 | Wu (2018), meta-analysis (32) |
|  | T18 | Hansen (2018), meta-analysis (22) |
|  | T32 | Petrelli (2019), meta-analysis (35) |
| **Eye** | T4 | Mattosinho (2019) meta-analysis (27) |
| **Head & Neck** | T4 | Gomez (2009), meta-analysis (37) |
|  | T1, T2, T4, T11 | Seoane (2012), meta-analysis (19) |
|  | T1, T2, T4, T11 | Seoane (2016), meta-analysis (18) |
|  | T29 | Lin (2016), meta-analysis (38) |
|  | T17, T30, T31 | Graboyes (2018), systematic review (23) |
| **Pediatric** | T4 | Brasme (2012), systematic review (28) |
|  | T4 | Lethaby (2013), systematic review (29) |
| **Prostate** | T18, T20 | van den Bergh (2014), systematic review (24) |
| **Ovarian** | T32 | Liu (2017), meta-analysis (46) |
|  | T32 | Usón (2017), meta-analysis (45) |
| **Multisite** | T1 - T9, T11 - T16, T18 - T27 | Neal (2015), systematic review (20) |
|  | T10 | Doubeni (2018), systematic review (30) |

<sup>a</sup> Lag time intervals correspond to those defined in Figure 2. Reviews on the same cancer site are sorted by publication year

### Findings from systematic reviews

The search strategy and characteristics of each included systematic review are detailed in **Supplementary Table 2**. We summarize in **Table 2** their overall characteristics and findings by cancer site and type as well as the corresponding lag time interval.

**Table 2.**
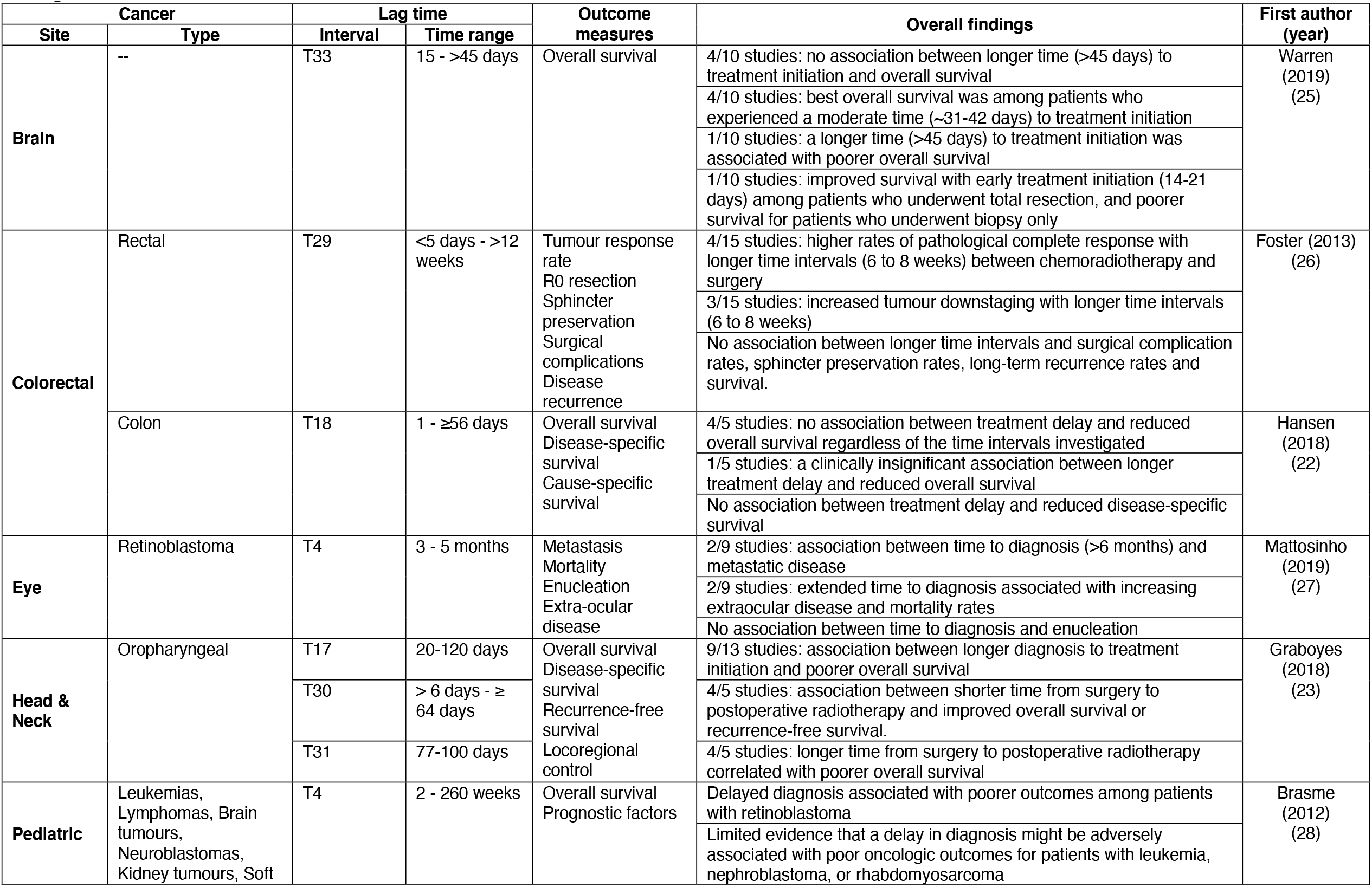
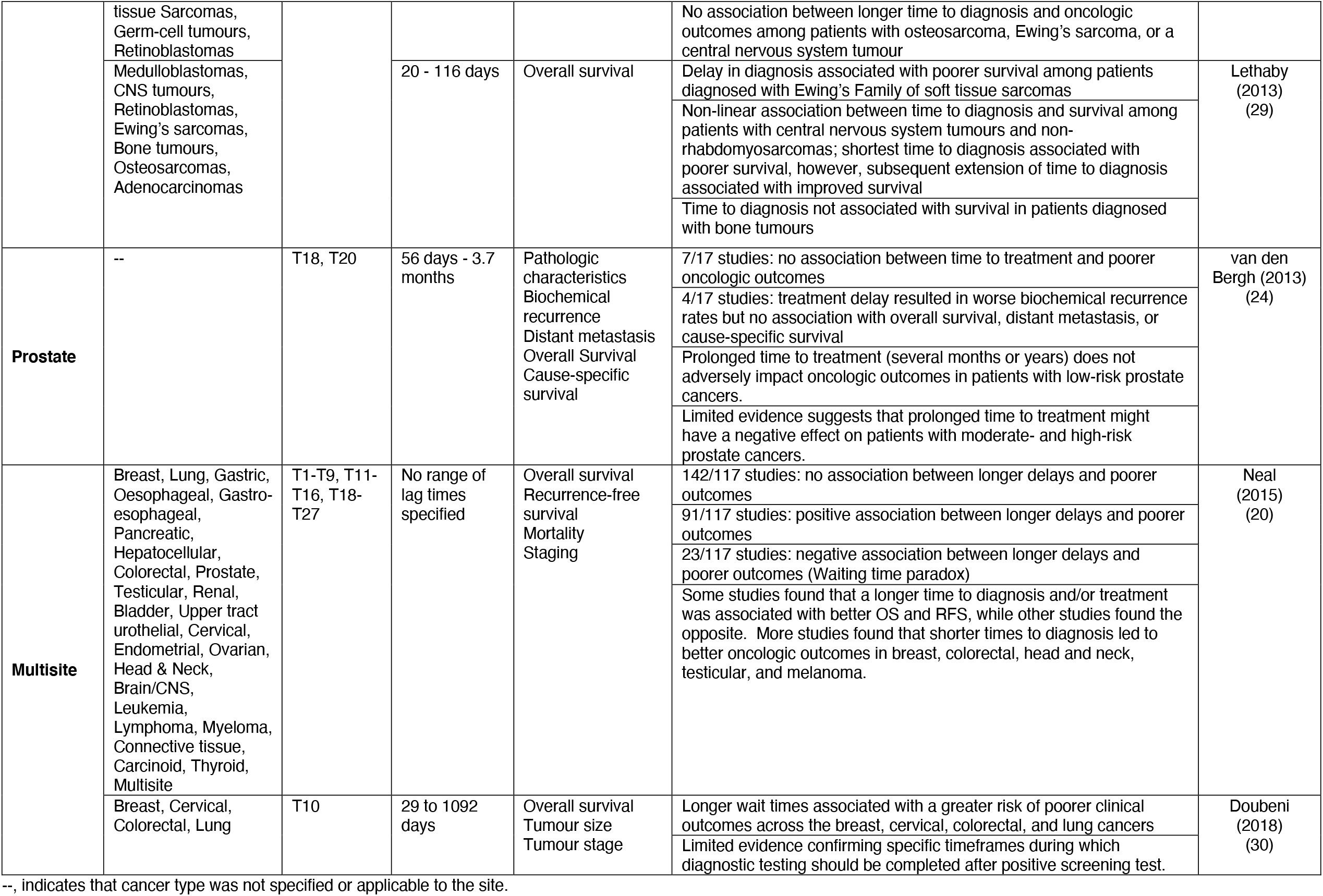
Characteristics of the systematic reviews on the association between time to cancer diagnosis and/or treatment and clinical outcomes, by cancer site/type and lag time interval

#### Brain

One systematic review investigating the impact of T33 (lag time between surgery and adjuvant chemotherapy) on overall survival (OS) among patients with brain cancer reported lack of consensus across included studies (25). Four of 10 included studies reported no association between T33 and OS among patients who experienced T33 > 45 days compared to those who experienced T33 < 45 days. A further four of 10 included studies reported improved OS among patients who experienced T33 between 31 and 42 days compared to those who experienced T33 < 31 days.

#### Colorectal

One review reported that higher pathologic complete response (pCR) rates (a prognostic measurement of treatment efficacy in the neoadjuvant setting) were associated with increased tumor downstaging among patients with rectal cancer experiencing time between neoadjuvant chemoradiotherapy (NACRT) and surgery (T29) > 6-8 weeks compared to those experiencing T29 < 6-8 weeks (26). In the same review, few included studies demonstrated that T29 > 6-8 weeks conferred higher pCR rates. With respect to the impact of prolonged time from diagnosis to surgery (T18) among patients with colon cancer, there was no association between extended T18, regardless of the length, and OS or disease-specific survival (DSS) (22).

#### Eye

Increased time between symptom onset and diagnosis (T4) was associated with increased rates of extraocular disease, metastatic disease, and mortality, but not enucleation, among patients with retinoblastomas (27).

#### Head & Neck

A systematic review on oropharyngeal cancers found extended time from diagnosis to surgery (T17) to be associated with poorer OS, shorter time from surgery to initiation of adjuvant radiotherapy (ART) (T30) to be associated with improved overall and recurrence-free survival (RFS), and prolonged time from surgery to completion of ART (T31) to be associated with poorer OS (23).

#### Pediatric

Worsened OS was reported among patients with retinoblastomas who experienced longer time between symptom onset and diagnosis (T4) (28). Among patients with Ewing’s sarcomas, poorer OS due to longer T4 was observed in one review (29), whereas no such association was reported in another review (28). While a non-linear association was reported between longer T4 and OS among patients with CNS tumours, where shorter T4 was associated with poorer OS and further extension of T4 was associated with improved OS (35), this association was not observed in the other review (28). Both reviews observed no association between extended T4 and OS among patients with osteosarcoma (28, 29).

#### Prostate

For patients with low-risk prostate cancers, extended time to treatment – either surgery (T18) or radiotherapy (T20) – was not associated with worsened oncologic outcomes or OS (24), however, some evidence suggests that these associations may be present among patients with moderate- and high-risk prostate cancers.

#### Multisite

Two reviews considered the impact of lag times in cancer care across multiple cancer sites (20, 30). One collected evidence on 25 different lag time intervals and 25 different cancer sites (19) and reported consensus across studies on likely associations between shorter times to diagnosis and improved oncologic outcomes among patients with breast, colorectal, head and neck, melanoma, and testicular cancers (20). The other review concluded that extended time from a positive screening test to diagnosis (T10) among patients with breast, cervical, colorectal, and lung cancers was associated with poorer oncological outcomes such as worsened OS and progressive tumor staging, however, this review emphasized lack of consensus regarding a particular timeframe during which diagnosis should be confirmed after a positive screening test as to mitigate these harmful outcomes (30).

#### Findings from meta-analyses

Detailed characteristics (i.e., search strategy, databases searched, number of hits, number of included studies, number of participants, etc.) of each included meta-analysis are presented in **Supplementary Table 3**. We summarize, by cancer site and type as well as lag time interval, their methodological characteristics and morbidity- (**Table 3**) and mortality- (**Table 4**) related findings.

**Table 3.**
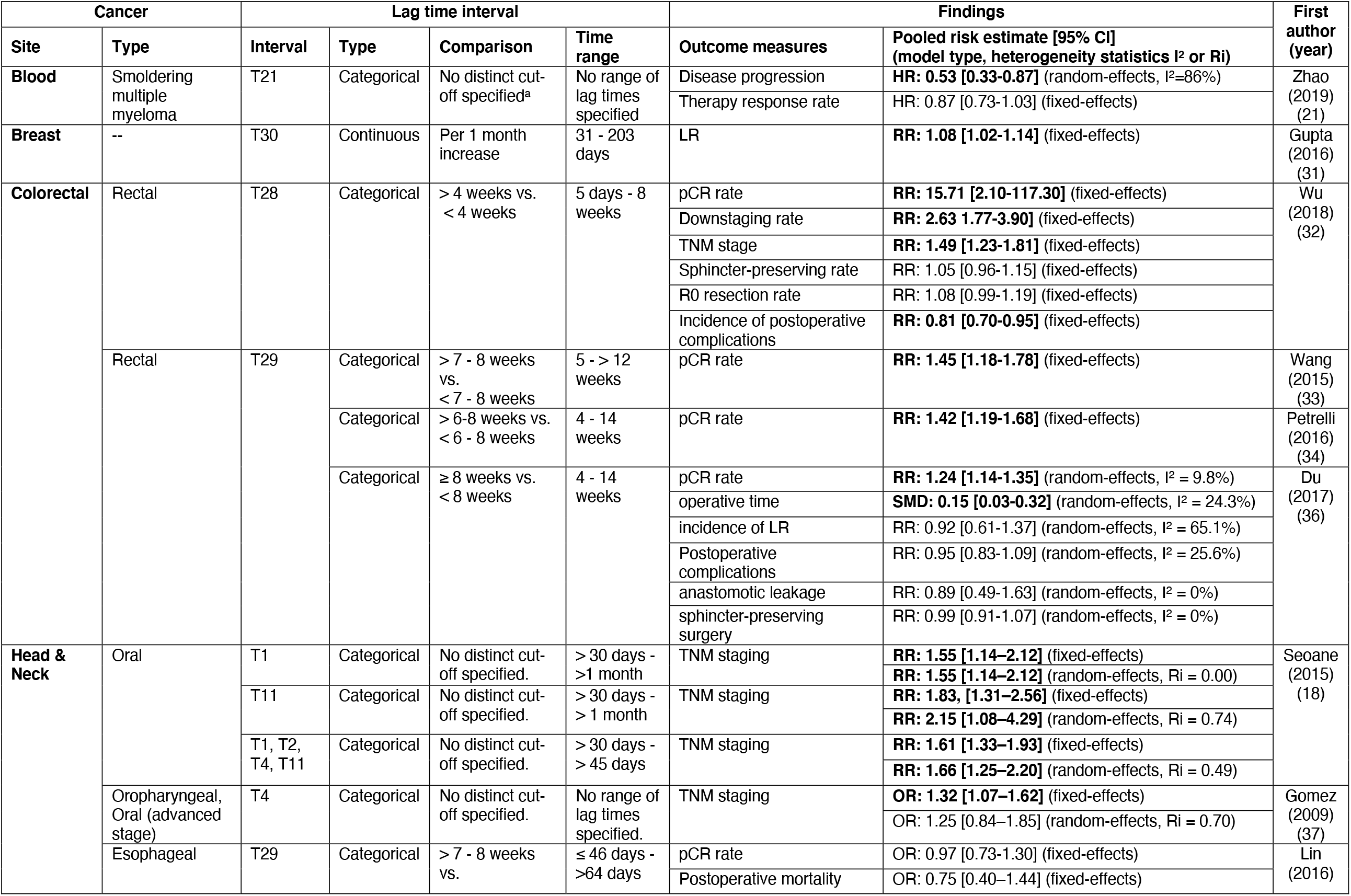
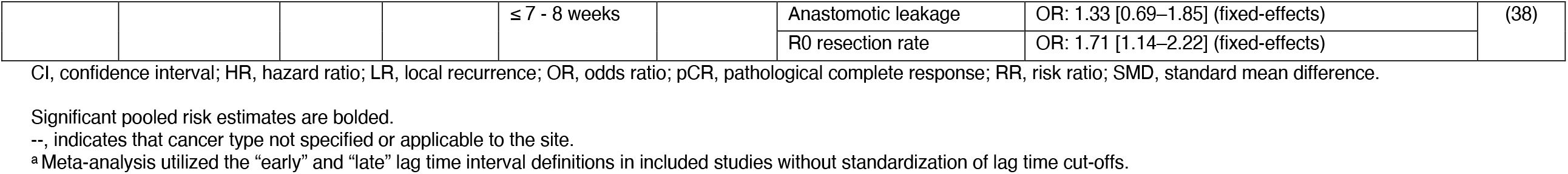
Morbidity-related findings of meta-analyses on the association between time to cancer diagnosis and/or treatment and clinical outcomes, by cancer site/type and lag time interval

**Table 4.**
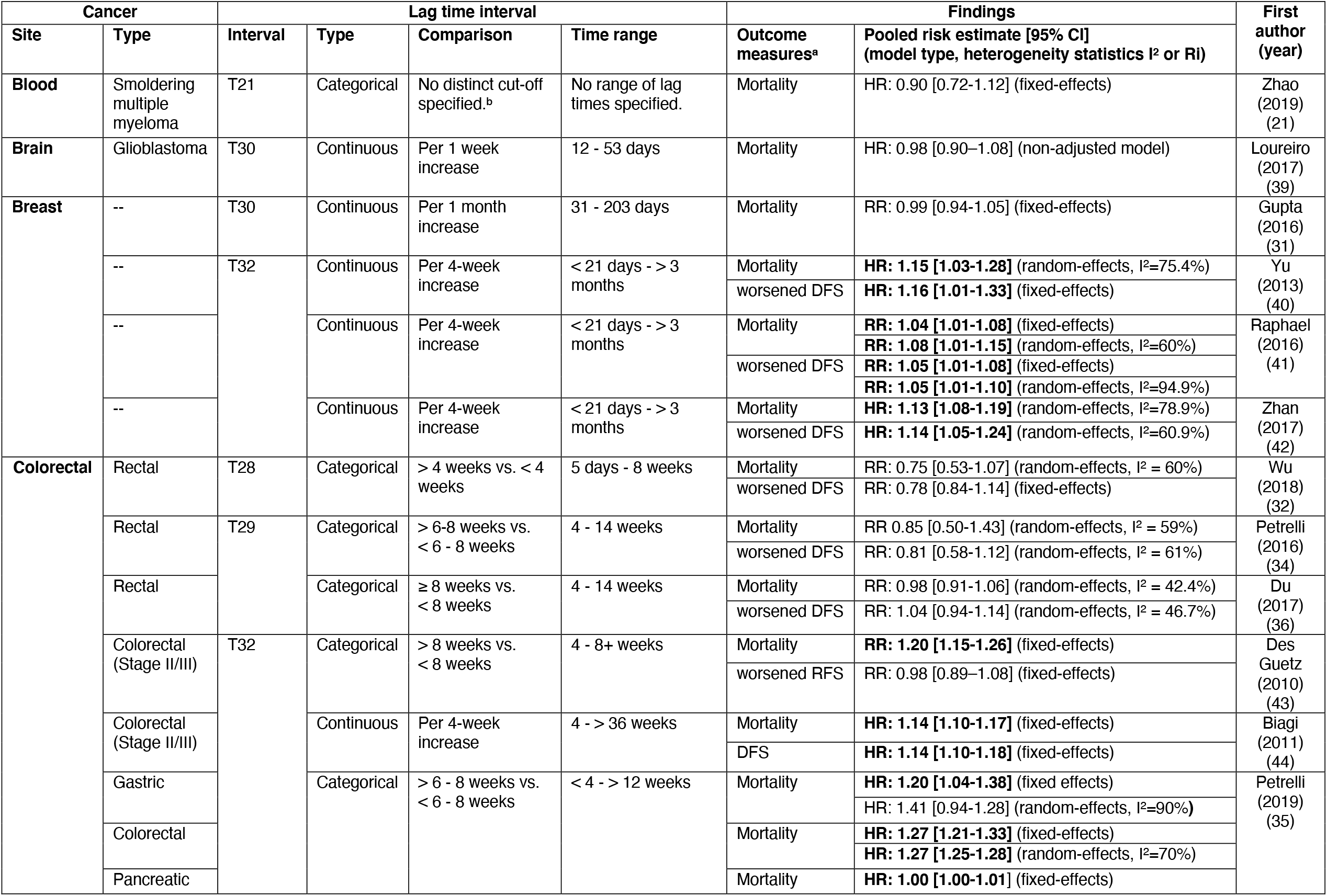
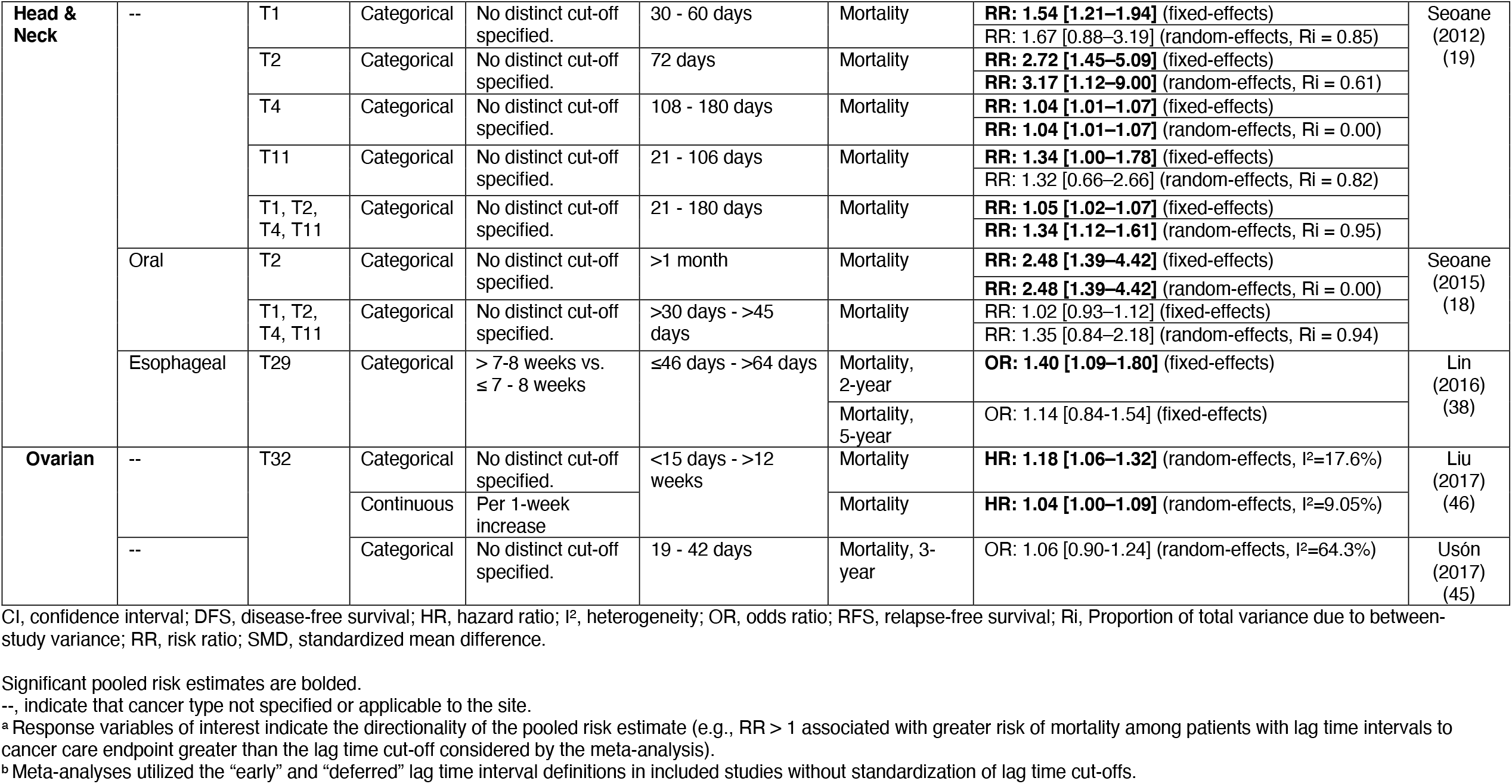
Mortality-related findings of meta-analyses on the association between time to cancer diagnosis and/or treatment and clinical outcomes, by cancer site/type and lag time interval.

### Morbidity-related findings

#### Blood

A significant association between shorter lag time between diagnosis and immunotherapy (T21) and decreased disease progression (HR: 0.53, 95% CI [0.33-0.87]) was found among patients with smoldering multiple myeloma (21).

#### Breast

A meta-analysis investigating the association between time between surgery and ART (T30) among patients with breast cancer reported a significant increase in risk of worsened locoregional control per 1-month increase of T30 (RR: 1.08, 95% CI [1.02-1.14]) (31).

#### Colorectal

pCR rate was the most common response variable investigated. Higher pCR rates were associated with time from neoadjuvant radiotherapy and surgery (T28) > 4 weeks (RR: 15.71, 95% CI [2.10-117.30]) (32), as well as with time from NACRT to surgery (T29) > 6 – 8 weeks (RR: 1.42, 95% CI [1.19-1.68]) (34), > 7 – 8 weeks (RR: 1.45, 95% CI [1.18-1.78]) (33), and ≥ 8 weeks (RR: 1.24, 95% CI [1.14-1.35] (36)).

#### Head & Neck

One meta-analysis investigated the association between time from symptom onset to [1] first being seen by a primary care provider (PCP) (T1), [2] referral to a specialist (T2), and [3] diagnosis (T4), as well as time from first being seen by a PCP to diagnosis (T11) on TNM staging among patients with oral cancer (18). There was greater risk of worsened TNM staging (based on “short” and “long” lag time cut-offs determined by included studies) associated with longer T1 (RR: 1.55, 95% CI [1.14–2.12]), T11 (RR: 2.15, 95% CI [1.08–4.29]), and any lag time (T1, T2, T4, or T11) (RR: 1.66 [1.25–2.20]). Another meta-analysis assessing the impact of time from symptom onset to diagnosis (T4) on TNM staging among patients with advanced-stage oral and oropharyngeal cancers found greater odds of increased TNM staging among patients who experienced T4 classified as “long” by included original studies (OR: 1.32, 95% CI [1.07–1.62]) (37).

### Mortality-related findings

#### Blood

No association was found between extended time from diagnosis to immunotherapy (T21) and risk of mortality among patients with smoldering multiple myelomas (21).

#### Brain

Time (per 1-week increase) between surgery and neoadjuvant radiotherapy (NART) (T30) was not associated with an increased risk of mortality among patients with brain cancers (39).

#### Breast

No change in risk of mortality was found per 1-month increase in the time between surgery and ART (T30) (31). Three meta-analyses reported a significant increase in risk of mortality per 1 month increase in T32 (time between surgery and adjuvant chemotherapy (ACT)) (HR: 1.15, 95% CI [1.03-1.28] (40); RR: 1.04, 95% CI [1.01-1.08] (41); and HR: 1.13, 95% CI [1.08-1.19] (42) as well as risk of worsened disease-free survival (DFS) per 1 month increase in T32 (HR: 1.16, 95% CI [1.01-1.33] (40); RR: 1.05, 95% CI [1.01-1.10] (41); HR: 1.14, 95% CI [1.05-1.24] (42)).

#### Colorectal

One meta-analysis investigated the impact of time between NART and surgery (T28) > 4 weeks on mortality and risk of worsened DFS and found no association (32). Two meta-analyses found no association between undergoing surgery > 6 – 8 weeks (34) or ≥ 8 weeks (36) after NACRT (T29) and risk of mortality as well as risk of worsened DFS. Three other meta-analyses evaluated the impact of time between surgery and ACT (T32) on mortality among patients with colorectal cancer (35, 43, 44). Those that classified the length of T32 categorically reported an increased risk of mortality among patients with colorectal cancer who experienced T32 > 6 – 8 weeks (HR: 1.27, 95% CI [1.25-1.28]) (35) and > 8 weeks (RR: 1.20, 95% CI [1.15-1.26]) (43). Similarly, a greater risk of mortality per 4-week increase in T32 was reported (HR: 1.14, 95% CI [1.10-1.17]) (44).

#### Head & Neck

Two meta-analyses investigated the influence of time from symptom onset to [1] first being seen by a PCP (T1), [2] referral for diagnosis (T2), [3] diagnosis (T4), and from first being seen by a PCP to diagnosis (T11), with one considering all head and neck cancers and the other oral cancers only (18, 19). For all head and neck cancers, significantly greater risks of mortality were found among patients who experienced extended T1 (RR: 1.54, 95% CI [1.21–1.94]), T2 (RR: 3.17, 95% CI [1.12–9.00]), T4 (RR: 1.04, 95% CI [1.01–1.07]), and T11 (RR: 1.34, 95% CI [1.00–1.78]) (19). Restriction to oral cancers revealed a greater risk of mortality among patients who experienced extended T2 (18). A meta-analysis investigating the effect of time from NACRT to surgery on mortality (T29) among patients with esophageal cancer found significantly greater odds of 2-year mortality among patients who experienced T29 > 7 – 8 weeks, with no association found between T29 > 7 – 8 weeks and the odds of 5-year mortality (38).

#### Ovarian

Of the two meta-analyses that evaluated the association between time from surgery to ACT (T32) and mortality among patients with ovarian cancer, one found a significantly greater risk of mortality per 1-week increase in T32 (HR: 1.04, 95% CI [1.00–1.09]) or among those who experienced extended T32 (HR: 1.18, 95% CI [1.06–1.32]) (46). The other meta-analysis reported no association between extended T32 and odds of 3-year mortality (45).

## DISCUSSION

We conducted a scoping review of systematic reviews and/or meta-analyses in order to characterize the body of pre-pandemic evidence on the known associations between lag time in cancer care and control and patient outcomes. Our comprehensive overview of the available peer-reviewed literature enabled the identification of either consistency or contradiction across reviews on the same lag time, cancer site, and outcome. Select comparisons across included meta-analyses investigating the same associations uncovered varying approaches to quantifying the effect of lag time on oncologic outcomes. Specifically, four lag time intervals for different cancer sites provided informative perspectives: [1] from surgery to ACT (T32) for breast cancer, [2] from NACRT to surgery (T29) for rectal cancer, [3] from surgery to ACT (T32) for colorectal cancer, and [4] from surgery to ACT (T32) for ovarian cancer. These comparisons revealed overarching methodological gaps in lag time literature.

### T32 and breast cancer

The three meta-analyses investigating the effect of extended time to ACT (T32) on mortality among patients with breast cancer reported a significantly greater risk of mortality per 1-month extension of T32 (40-42). Although they provided consistent conclusions, differences in the magnitude of the reported risk estimates could be attributed to conflicting inclusion of a registry-based study (47) that found a significant association between extended T32 and mortality. This registry-based study (47) was included in Yu et al.’s (40) and Zhan et al.’s (42) meta-analyses, which yielded risk estimates greatest in magnitude, and accounted for 21.24% and 8.09% of the weight in their analyses, respectively. Raphael et al. (41), who reported the most conservative pooled risk estimate, did not include this registry-based study (47). Raphael et al. (41) argued that inclusion of this registry-based study in Yu et al.’s analysis (40) could have introduced bias by confounding or misclassification of ACT as palliative rather than curative, resulting in an overestimation of risk associated with prolonged T32. By extension, this argument could also apply to Zhan et al.’s analysis (42). However, Raphael et al.’s argument relied on the assumption that those who received palliative ACT were not only less likely to survive, but also more likely to experience longer time to ACT after surgery, thus inducing an artifactual association between shorter time from surgery to ACT and mortality. Because it is unknown whether patients undergoing palliative care would be considered lower priority for receiving ACT or, conversely, experience shorter time to ACT due to directed resources specific to palliative care units, it is unclear as to whether the possible inclusion of patients undergoing palliative ACT in the registry-based study (47) would have led to an over- or under-estimation of Yu et al.’s (40) and Zhan et al.’s (42) risk estimates.

### T29 and colorectal cancer

The three meta-analyses evaluating the impact of time from NACRT to surgery (T29) on pCR rates among patients with advanced rectal cancer provided different conclusions regarding the optimal timing of surgery (33, 34, 36). Each reported protective associations of extended T29 for pCR rates using different cut-offs to define “longer” versus “control” T29: 7 weeks (33), 6-8 weeks (34), and 8 weeks after NACRT (36). These cut-offs were determined based on the seminal Lyon R90-01 trial, which investigated the impact of 6-8 weeks between NART and sphincter-preserving surgery compared to 2 weeks on pathological downstaging among patients with rectal cancer and established 6-8 weeks as an accepted lag time of NART or NACRT after surgery in clinical practice (48). However, an evaluation of a 6-8 week length of time between preoperative radiotherapy and surgery in the trial might not have precluded the potential for shorter time between preoperative chemoradiotherapy and surgery (T29) (> 2 weeks and < 6-8 weeks), or longer T29 (> 6-8 weeks) to sustain benefit for patients with rectal cancer.

Considering this uncertainty, Wang et al. conducted subgroup analyses based on different cut-offs of T29 (5, 6, 7, 8, 10, and 12 weeks) (**Supplemental Table 3**) and found that performing surgery at 7 and 8 weeks yielded a significantly improved pCR rates compared to performing surgery earlier, at 5 or 6 weeks, or later, at 10 or 12 weeks (33). This reported “optimal window”, which is narrower than the clinically accepted 6-8 week window, suggests that benefits associated with NACRT for patients with advanced rectal cancer are dependent on the timing of surgery and also on inter-individual variation in response to NACRT. Among rectal cancer patients, surgery performed too soon may not allow for the maximal anti-tumorigenic response to NACRT, however, waiting too long could mitigate any potential benefit that could be maintained from NACRT and permit tumor repopulation.

### T32 and colorectal cancer

Three meta-analyses (35, 43, 44) investigating the association between time from surgery to ACT on mortality among patients with colorectal cancer reported significant associations between extended time from surgery to ACT (T32) and risk of mortality. Yet, all three (35, 43, 44) used differing statistical methods with regards to lag time variable classification. Des Guetz et al.’s (43) and Petrelli et al.’s (2019) (35) considered lag time categorically with respective cut-offs of > 8 weeks and > 6-8 weeks, while Biagi et al. (44) considered lag time as a continuous variable. Biagi et al. (44) excluded four studies (49-52) that were included in Des Guetz et al.’s analysis (41) based on lack of adjustment for potential confounders. Two studies (53, 54) included in Biagi et al.’s (44) and Des Guetz et al.’s (43) analysis, which cumulatively contributed 69.05% of the weight in Biagi et al.’s analysis (weight was not reported by Des Guetz et al.), reported risk estimates highly similar to those yielded from the two meta-analyses. After exclusion of the two largest studies (53, 54) in both analyses, significance was maintained (**Supplemental Table 3**).

### T32 and ovarian cancer

The two meta-analyses on the association between time to ACT after surgery (T32) and risk of mortality among patients with ovarian cancer reported contradictory findings; one found an increased risk of mortality due to longer T32 (46), but not the other (45). Key methodological differences could account for these conflicting findings. Although there was substantial overlap of included studies between the meta-analyses, the one that found an increased risk of mortality due to longer T32 (46) stratified the analysis by lag time variable type (categorical or continuous), while the other meta-analysis that found no association (45) included studies independent of the lag time variable type. Inclusion of studies in the same analysis independent of or dependent upon how lag time was considered likely contributed to the substantial heterogeneity observed in the meta-analysis that did not stratify by lag time variable type (I^2^ = 64.3%) (45) compared to the one that did (I^2^ = 17.6%, 9.05%) (46). Notably, the cut-offs defining “early” versus “deferred” lag time intervals in both meta-analyses were based on those used in the included original studies. It is also likely that the wide range of lag time intervals across studies might have influenced the precision and significance of the reported risk estimates, hence the importance of standardization of lag time cut-offs in meta-analyses and the impact of lag time variable type on statistical findings.

### Methodological gaps in lag time literature

In juxtaposing the design and analytical approaches of included reviews, we identified three predominant methodological gaps in lag time literature. Firstly, consideration of change in intervention modality over time was inconsistent across included reviews. Failure to account for improvement of clinical protocols and treatment regimens, which can confer greater protection against morbidity- and mortality-related outcomes, could have biased the observed risk estimates based on recency of included studies and their corresponding weight in the meta-analyses. For example, the three meta-analyses (40-42) that reported significant associations between time to ACT after surgery (T32) and mortality among patients with breast cancer included studies for which chemotherapy regimens were anthracycline-based and/or CMF regimens with the earliest included study dating to 1989 (55). This could constrain generalizability of results to patients treated with more recent ACT regimens which primarily include taxanes as standard care (56). It is noteworthy, however, that apart from the recent advent of immunotherapy and targeted therapies, which fall under the umbrella of precision medicine, there have not been major paradigm shifts in treatment provision during the timespan covered by our search (1 January 2010 to 31 December 2019). Nevertheless, our restriction on publication date of included systematic reviews and/or meta-analyses does not prevent the inclusion of studies that evaluate interventions that have since evolved in type, dosing, and/or timing. Indeed, future ubiquity of novel therapies under the purview of precision medicine will necessitate consideration of the evolution of cancer care in meta-analytical lag time research. In such cases, conducting sensitivity analyses based on treatment type, regimen, or date of publication can provide a clearer understanding of the external validity of reported risk estimates. The findings of select included meta-analyses that did conduct such sensitivity analyses (21, 42, 46) (**Supplemental Table 3**) demonstrated the variation in significance of the association between lag time and clinical outcomes due to treatment specification and recency of publication in addition to baseline patient characteristics, such as demographics, stage, date of administration, dosing, type of therapy, and clinical decisions.

Secondly, ambiguity in defining lag time interval start and endpoints in the reviews and original studies could lead to exposure misclassification. For example, in the case of categorical lag time variables, specification of whether an endpoint is considered as initiation of NACRT, completion of NACRT, or time of medical charting of NACRT, could influence the length of the lag time interval measured and classification of such exposure as experimental or control.

The Aarhus statement, a set of recommendations and checklist resulting from discussion regarding methodological concerns in research conducted on lag times to cancer diagnosis, identified unclear definitions of start and endpoints in lag times, as well as inconsistency in defining these points across studies evaluating the same lag time as primary obstacles to aggregating research on lag time to cancer diagnosis (57). We argue that the same concerns would apply to research on lag time between any start and endpoint on the cancer care continuum, not only within time to cancer diagnosis.

Some of the included qualitative systematic reviews initially intended to meta-analyze the abstracted data (22, 23, 26, 29), however, variability in start and endpoint definitions of lag times across included studies and in type of lag time variable (categorical versus continuous) prevented researchers from pursuing quantitative analyses. This lack of clarity of exposure definitions stymies lag time research and calls for an expansion of the pre-existing Aarhus statement (57) to encompass lag time research across the entire cancer care continuum. Refinements of the Aarhus statement integrating suggestions from clinicians and cancer diagnosis researchers have been proposed (58), yet are still restricted to cancer diagnosis, rather than applicable to the entire cancer care continuum. Additions to the Aarhus statement regarding the entire continuum could provide not only recommendations for standardizing lag time definitions in randomized-controlled trials and observational studies, but also for justifying classification of lag time categorically versus continuously (e.g., evaluation of a lag time cut-off in standard care). Apart from residual confounding that can arise from categorical classification of lag time, the method of defining lag time categories using cut-offs can substantially impact resulting risk estimates and their corresponding uncertainty. Some included meta-analyses (18, 19, 21, 37, 45, 46) utilized categories of “early” and “deferred” care defined by original studies. Even when classifying lag time categorically is appropriate, not standardizing cut-offs across included studies can give rise to more significant variation in risk estimates and constrict interpretation of resulting findings. Overall, recommendations for defining lag time exposure across the entire continuum and strategizing statistical classification of lag time exposure is necessary for improving the quality and utility of future lag time research.

Thirdly, confounding by indication needs to be taken into consideration in research conducted on lag times in cancer care. Otherwise monikered the “waiting-time paradox”, confounding by indication is the implication that patients with more advanced stage disease are prioritized for referral and subsequent treatment within a given health system more rapidly than patients with early-stage, less severe disease. The overall effect of this prioritization leads to the indication that early referral and/or treatment is associated with higher mortality; this survival trend is in direct contradiction with the typical log-linear approach taken in meta-analyses on the association between extended lag time to cancer treatment and risk of mortality. The presence of the waiting-time paradox in meta-analyses can be most clearly identified when lag time is treated as a continuous rather than categorical variable. However, this can be difficult as studies included in meta-analyses often regard lag times as dichotomous exposure. Few of the meta-analyses included in the current scoping review reported risk estimates for lag time intervals as a continuous exposure (31, 39-42, 44, 46). Notwithstanding appropriate justification for treating lag time categorically, either sensitivity analyses using different cut-offs of categorical lag time or meta-analyses using continuous lag time could facilitate identification of the waiting-time paradox and provide insights regarding its mechanisms.

Apart from inherent limitations of the included systematic reviews and meta-analyses, some overarching limitations in the methodology of our scoping review need to be acknowledged. Restricting our search to systematic reviews and/or meta-analyses published after 2010 did not prevent the introduction of medical interventions that have changed over time in included literature, however, our reporting of stratified analyses performed in included meta-analyses, when available, by treatment specifications (e.g., dosing, chemotherapy type) does provide context regarding modification of lag time on clinical outcomes by intervention modality. Further, even though some meta-analyses evaluated the same parameters, variability in statistical methods limited the scope of our comparisons between these meta-analyses. While we did not conduct quality assessment due to the nature of the included literature, we did describe characteristics of each review which could aid in assessing the validity and generalizability of each review’s findings. Finally, despite the importance of examining treatment variations among patients within clinical trial settings, which are circumscribed to well-established rules and procedures, real-world evidence, such as data from electronic medical or health records, can provide further insight into patient profiles, treatment choice, drug adherence, and adverse event management. These individual-level factors, which are not captured by the included meta-analyses, can appreciably influence oncologic outcomes and thus the true causal effect of lag time on morbidity- and mortality-related outcomes.

### Contextualizing findings of the scoping review amidst the COVID-19 pandemic

Our comprehensive map of lag time intervals and clinical outcomes across multiple cancer sites is intended to serve as a reference point for future research evaluating the pandemic’s impact on lag times in cancer control and care. As it can take years for cancer-related survival outcomes to accrue, it is too soon to accurately quantify the impact of extended times to diagnosis and treatment attributable to the pandemic. Hence, the perspectives presented herein on the impact of lag time in cancer control and care not only aid in benchmarking pandemic-induced lag time compared to standard-of-care lag time experienced prior to the pandemic, but also inform ongoing research on these unprecedented lag times experienced by patients.

Recently published modelling studies simulating the long-term impact of pandemic-induced lag times on cancer-related outcomes have informed the degree of tolerability of the widespread consequences of the pandemic on cancer care systems (2, 13). A U.K. population-based modelling study predicting the impact of lag times to diagnosis of colorectal, breast, lung, and esophageal cancers on survival during the 12 months after national lockdown measures began estimated increases of 7.9-9.6%, 15.3-16.6%, 4.8-5.3%, and 5.8-6.0% in deaths due to breast, colorectal, lung, and esophageal cancers, respectively, up to 5 years after diagnosis compared to pre-pandemic data (2). Another modelling study from our group projected a 2% increase in cancer deaths, or an excess 355,172 life-years-lost, in Canada between 2020 and 2030 due to pandemic-induced lag times to both cancer diagnosis and treatment, assuming the cancer care system returned to normal capacity in 2021 (15). Triage systems prioritizing patients into treatment and diagnostic systems (7), on top of system-related constraints, have resulted in unprecedented lag times that exceed those experienced pre-pandemic (6). This evokes the question as to whether adverse consequences associated with such pandemic-related lag-times will be harsher than those described in retrospective data.

The prevailing concern is that any attenuation of the impact of pandemic-induced lag time on cancer diagnosis and treatment is dependent on immediate recovery of cancer care infrastructure. As was seen most recently with the Omicron variant, which stressed already over-burdened health care systems worldwide, healthcare systems’ recovery to full capacity is tenuous. Such fragility signals for an ongoing need to quantify and contextualize lag time’s impact on cancer-related outcomes. This need was highlighted by Hanna et al.’s meta-analysis, which reported generalizable measures of effect of four-week-lag time to treatment, stratified by modality, across seven common cancers (59). While meta-analyses, especially the one by Hanna et al. (59), can serve as tools for parameterizing models (15) or summarizing the impact of time to treatment across common cancers (60), they may not capture information relevant to particular populations and outcomes. Three included meta-analyses (33, 34, 36), which reported improved pCR rates among patients with advanced-stage rectal cancer who experienced time between NACRT and surgery of 6-8 weeks, demonstrated the biological benefit of lag time within the context of the intervention. Similarly, extended lag time to subsequent steps in care can be clinically appropriate with regards to patient rehabilitation post-treatment. The impact of lag time to diagnosis and/or treatment may also vary across different types of cancer with differing risk or rates of development within the same site (e.g., breast cancer). That is, just as prolonged lag time can be deleterious, which is often how it is connoted, it can also denote advantageous clinical characteristics. Our scoping review aimed to attend to the same need of summarizing lag time’s impact on oncologic outcomes, however, with the intention of preserving the granularity of these associations across multiple cancer sites, with clearly mapped lag time interval start and endpoints. Our tracing of defined lag times across relevant systematic reviews and/or meta-analyses can serve as a pre-pandemic reference when assessing the tolerability of pandemic-induced lag times. It could also lay the groundwork for observational and meta-analytical research on lag time intervals’ influence on oncologic outcomes across sites beyond the most common ones, such as breast, lung, and colorectal.

## CONCLUSION

Through the aggregation of known associations between lag time and oncologic outcomes and exploration into gaps in lag time research, this scoping review can guide future studies and meta-analyses in the discipline. Our lag time interval timeline, or mapping of lag times on the cancer care continuum, emphasizes the granularity of exposure classification in cancer care. This timeline can act as a blueprint for future studies assessing lag time intervals and/or multiple cancer sites. With regards to the COVID-19 pandemic, our extensive characterization of the effect of lag time on oncologic outcomes could aid in gauging lag times in cancer care experienced during the pandemic.

## Data Availability

All data from included studies presented by the current work are available on MEDLINE, EMBASE, Web of Science, and/or Cochrane Library of Systematic Reviews. We include our systematic search strategy in Supplementary Material.

## CONTRIBUTORS

All authors contributed to the conception and design of this review. EF and RA designed the search strategy. PT, EF, and RA conducted the electronic search, retrieved and screened articles for eligibility, and extracted relevant data. PT synthesized the abstracted data, produced all tables and figures, and drafted the manuscript under the supervision of MZ and ELF. All authors revised and approved the final submitted manuscript.

## References

1. Mast C, Munoz del Rio A. Delayed Cancer Screenings – A Second Look. Report. Epic Health Research Network: Epic Research; 2020.

2. Maringe C, Spicer J, Morris M, Purushotham A, Nolte E, Sullivan R, Rachet B, Aggarwal A. The impact of the COVID-19 pandemic on cancer deaths due to delays in diagnosis in England, UK: a national, population-based, modelling study. Lancet Oncol. 2020;21(8):1023–3410.1016/S1470-2045(20)30388-0.

3. Villain P, Carvalho AL, Lucas E, Mosquera I, Zhang L, Muwonge R, Selmouni F, Sauvaget C, Basu P. Cross-sectional survey of the impact of the COVID-19 pandemic on cancer screening programs in selected low-and middle-income countries: Study from the IARC COVID-19 impact study group. Int J Cancer. 2021;149(1):97–10710.1002/ijc.33500.

4. Walker MJ, Wang J, Mazuryk J, Skinner S-M, Meggetto O, Ashu E, Habbous S, Nazeri Rad N, Espino-Hernández G, Wood R, Chaudhry M, Vahid S, Gao J, Gallo-Hershberg D, Gutierrez E, Zanchetta C, Langer D, Zwicker V, Rey M, Tammemägi MC, Tinmouth J, Kupets R, Chiarelli AM, Singh S, Warde P, Forbes L, Dobranowski J, Irish J, Rabeneck L, Group CCOC-IW. Delivery of Cancer Care in Ontario, Canada, During the First Year of the COVID-19 Pandemic. JAMA Network Open. 2022;5(4):e228855–e10.1001/jamanetworkopen.2022.8855.

5. Gotlib Conn L, Tahmasebi H, Meti N, Wright FC, Thawer A, Cheung M, Singh S. Cancer Treatment During COVID-19: A Qualitative Analysis of Patient-Perceived Risks and Experiences with Virtual Care. J Patient Exp. 2021;8:23743735211039328 10.1177/23743735211039328.

6. Jazieh AR, Akbulut H, Curigliano G, Rogado A, Alsharm AA, Razis ED, Mula-Hussain L, Errihani H, Khattak A, Guzman RBD, Mathias C, Alkaiyat MOF, Jradi H, Rolfo C, Care obotIRNoC-IoC. Impact of the COVID-19 Pandemic on Cancer Care: A Global Collaborative Study. JCO Glob Oncol. 2020(6):1428–38 10.1200/go.20.00351.

7. Farah E, Ali R, Tope P, El-Zein M, Franco EL, McGill Task Force on COVID-19 and Cancer. A Review of Canadian Cancer-Related Clinical Practice Guidelines and Resources during the COVID-19 Pandemic. Curr Oncol. 2021;28(2):1020–33 10.3390/curroncol28020100.

8. Carvalho AS, Fernandes ÓB, de Lange M, Lingsma H, Klazinga N, Kringos D. Changes in the quality of cancer care as assessed through performance indicators during the first wave of the COVID-19 pandemic in 2020: a Scoping Review. medRxiv. 2022:2022.02.23.22271303 10.1101/2022.02.23.22271303.

9. Teckie S, Andrews JZ, Chen WC-Y, Goenka A, Koffler D, Adair N, Potters L. Impact of the COVID-19 Pandemic Surge on Radiation Treatment: Report From a Multicenter New York Area Institution. J Oncol Pract. 2021;17(9):e1270–e7 10.1200/op.20.00619.

10. Elkrief A, Kazandjian S, Bouganim N. Changes in Lung Cancer Treatment as a Result of the Coronavirus Disease 2019 Pandemic. JAMA Oncol. 2020;6(11):1805–6 10.1001/jamaoncol.2020.4408.

11. Moletta L, Pierobon ES, Capovilla G, Costantini M, Salvador R, Merigliano S, Valmasoni M. International guidelines and recommendations for surgery during Covid-19 pandemic: A Systematic Review. Int J Surg. 2020;79:180–810.1016/j.ijsu.2020.05.061.

12. Sharpless NE. COVID-19 and cancer. Science. 2020;368(6497):1290 doi:10.1126/science.abd3377.

13. Sud A, Torr B, Jones ME, Broggio J, Scott S, Loveday C, Garrett A, Gronthoud F, Nicol DL, Jhanji S, Boyce SA, Williams M, Riboli E, Muller DC, Kipps E, Larkin J, Navani N, Swanton C, Lyratzopoulos G, McFerran E, Lawler M, Houlston R, Turnbull C. Effect of delays in the 2-week-wait cancer referral pathway during the COVID-19 pandemic on cancer survival in the UK: a modelling study. Lancet Oncol. 2020;21(8):1035–4410.1016/s1470-2045(20)30392-2.

14. Burger EA, Jansen EE, Killen J, Kok IM, Smith MA, Sy S, Dunnewind N, N GC, Haas JS, Kobrin S, Kamineni A, Canfell K, Kim JJ. Impact of COVID-19-related care disruptions on cervical cancer screening in the United States. J Med Screen. 2021;28(2):213–6 10.1177/09691413211001097.

15. Malagón T, Yong JHE, Tope P, Miller Jr. WH, Franco EL. Predicted long-term impact of COVID-19 pandemic-related care delays on cancer mortality in Canada. Int J Cancer. 2022;150(8):1244–5410.1002/ijc.33884.

16. Tricco AC, Lillie E, Zarin W, O’Brien KK, Colquhoun H, Levac D, Moher D, Peters MDJ, Horsley T, Weeks L, Hempel S, Akl EA, Chang C, McGowan J, Stewart L, Hartling L, Aldcroft A, Wilson MG, Garritty C, Lewin S, Godfrey CM, Macdonald MT, Langlois EV, Soares-Weiser K, Moriarty J, Clifford T, Tunçalp Ö, Straus SE. PRISMA Extension for Scoping Reviews (PRISMA-ScR): Checklist and Explanation. Ann Intern Med. 2018;169(7):467–73 10.7326/M18-0850.

17. Ouzzani M, Hammady H, Fedorowicz Z, Elmagarmid A. Rayyan—a web and mobile app for systematic reviews. Systematic Reviews. 2016;5(1):210 10.1186/s13643-016-0384-4.

18. Seoane J, Alvarez-Novoa P, Gomez I, Takkouche B, Diz P, Warnakulasiruya S, Seoane-Romero JM, Varela-Centelles P. Early oral cancer diagnosis: The Aarhus statement perspective. A systematic review and meta-analysis. Head Neck. 2016;38 Suppl 1:E2182–9 10.1002/hed.24050.

19. Seoane J, Takkouche B, Varela-Centelles P, Tomás I, Seoane-Romero JM. Impact of delay in diagnosis on survival to head and neck carcinomas: a systematic review with meta-analysis. Clin Otolaryngol. 2012;37(2):99–106 10.1111/j.1749-4486.2012.02464.x.

20. Neal RD, Tharmanathan P, France B, Din NU, Cotton S, Fallon-Ferguson J, Hamilton W, Hendry A, Hendry M, Lewis R, Macleod U, Mitchell ED, Pickett M, Rai T, Shaw K, Stuart N, Tørring ML, Wilkinson C, Williams B, Williams N, Emery J. Is increased time to diagnosis and treatment in symptomatic cancer associated with poorer outcomes? Systematic review. Br J Cancer. 2015;112 Suppl 1(Suppl 1):S92–107 10.1038/bjc.2015.48.

21. Zhao AL, Shen KN, Wang JN, Huo LQ, Li J, Cao XX. Early or deferred treatment of smoldering multiple myeloma: a meta-analysis on randomized controlled studies. Cancer Manag Res. 2019;11:5599–611 10.2147/cmar.S205623.

22. Hangaard Hansen C, Gögenur M, Tvilling Madsen M, Gögenur I. The effect of time from diagnosis to surgery on oncological outcomes in patients undergoing surgery for colon cancer: A systematic review. Eur J Surg Oncol. 2018;44(10):1479–85 10.1016/j.ejso.2018.06.015.

23. Graboyes EM, Kompelli AR, Neskey DM, Brennan E, Nguyen S, Sterba KR, Warren GW, Hughes-Halbert C, Nussenbaum B, Day TA. Association of Treatment Delays With Survival for Patients With Head and Neck Cancer: A Systematic Review. JAMA Otolaryngol Head Neck Surg. 2019;145(2):166–77 10.1001/jamaoto.2018.2716.

24. van den Bergh RC, Albertsen PC, Bangma CH, Freedland SJ, Graefen M, Vickers A, van der Poel HG. Timing of curative treatment for prostate cancer: a systematic review. Eur Urol. 2013;64(2):204–15 10.1016/j.eururo.2013.02.024.

25. Warren KT, Liu L, Liu Y, Milano MT, Walter KA. The Impact of Timing of Concurrent Chemoradiation in Patients With High-Grade Glioma in the Era of the Stupp Protocol. Front Oncol. 2019;9(186) 10.3389/fonc.2019.00186.

26. Foster JD, Jones EL, Falk S, Cooper EJ, Francis NK. Timing of surgery after long-course neoadjuvant chemoradiotherapy for rectal cancer: a systematic review of the literature. Dis Colon Rectum. 2013;56(7):921–30 10.1097/DCR.0b013e31828aedcb.

27. Mattosinho CCS, Moura A, Oigman G, Ferman SE, Grigorovski N. Time to diagnosis of retinoblastoma in Latin America: A systematic review. Pediatr Hematol Oncol. 2019;36(2):55–72 10.1080/08880018.2019.1605432.

28. Brasme JF, Morfouace M, Grill J, Martinot A, Amalberti R, Bons-Letouzey C, Chalumeau M. Delays in diagnosis of paediatric cancers: a systematic review and comparison with expert testimony in lawsuits. Lancet Oncol. 2012;13(10):e445–59 10.1016/s1470-2045(12)70361-3.

29. Lethaby CD, Picton S, Kinsey SE, Phillips R, van Laar M, Feltbower RG. A systematic review of time to diagnosis in children and young adults with cancer. Arch Dis Child. 2013;98(5):349–55 10.1136/archdischild-2012-303034.

30. Doubeni CA, Gabler NB, Wheeler CM, McCarthy AM, Castle PE, Halm EA, Schnall MD, Skinner CS, Tosteson ANA, Weaver DL, Vachani A, Mehta SJ, Rendle KA, Fedewa SA, Corley DA, Armstrong K. Timely follow-up of positive cancer screening results: A systematic review and recommendations from the PROSPR Consortium. CA Cancer J Clin. 2018;68(3):199–216 10.3322/caac.21452.

31. Gupta S, King WD, Korzeniowski M, Wallace DL, Mackillop WJ. The Effect of Waiting Times for Postoperative Radiotherapy on Outcomes for Women Receiving Partial Mastectomy for Breast Cancer: a Systematic Review and Meta-Analysis. Clin Oncol (R Coll Radiol). 2016;28(12):739–49 10.1016/j.clon.2016.07.010.

32. Wu H, Fang C, Huang L, Fan C, Wang C, Yang L, Li Y, Zhou Z. Short-course radiotherapy with immediate or delayed surgery in rectal cancer: A meta-analysis. Int J Surg. 2018;56:195–202 10.1016/j.ijsu.2018.05.031.

33. Wang XJ, Zheng ZR, Chi P, Lin HM, Lu XR, Huang Y. Effect of Interval between Neoadjuvant Chemoradiotherapy and Surgery on Oncological Outcome for Rectal Cancer: A Systematic Review and Meta-Analysis. Gastroenterol Res Pract. 2016;2016:6756859 10.1155/2016/6756859.

34. Petrelli F, Sgroi G, Sarti E, Barni S. Increasing the Interval Between Neoadjuvant Chemoradiotherapy and Surgery in Rectal Cancer: A Meta-analysis of Published Studies. Ann Surg. 2016;263(3):458–64 10.1097/sla.0000000000000368.

35. Petrelli F, Zaniboni A, Ghidini A, Ghidini M, Turati L, Pizzo C, Ratti M, Libertini M, Tomasello G. Timing of Adjuvant Chemotherapy and Survival in Colorectal, Gastric, and Pancreatic Cancer. A Systematic Review and Meta-Analysis. Cancers (Basel). 2019;11(4) 10.3390/cancers11040550.

36. Du D, Su Z, Wang D, Liu W, Wei Z. Optimal Interval to Surgery After Neoadjuvant Chemoradiotherapy in Rectal Cancer: A Systematic Review and Meta-analysis. Clin Colorectal Cancer. 2018;17(1):13–24 10.1016/j.clcc.2017.10.012.

37. Gómez I, Seoane J, Varela-Centelles P, Diz P, Takkouche B. Is diagnostic delay related to advanced-stage oral cancer? A meta-analysis. Eur J Oral Sci. 2009;117(5):541–6 10.1111/j.1600-0722.2009.00672.x.

38. Lin G, Han SY, Xu YP, Mao WM. Increasing the interval between neoadjuvant chemoradiotherapy and surgery in esophageal cancer: a meta-analysis of published studies. Dis Esophagus. 2016;29(8):1107–14 10.1111/dote.12432.

39. Loureiro LVM, Victor EdS, Callegaro-Filho D, Koch LdO, Pontes LdB, Weltman E, Rother ET, Malheiros SMF. Minimizing the uncertainties regarding the effects of delaying radiotherapy for Glioblastoma: A systematic review and meta-analysis. Radiother Oncol. 2016;118(1):1–8 10.1016/j.radonc.2015.11.021.

40. Yu KD, Huang S, Zhang JX, Liu GY, Shao ZM. Association between delayed initiation of adjuvant CMF or anthracycline-based chemotherapy and survival in breast cancer: a systematic review and meta-analysis. BMC Cancer. 2013;13:240 10.1186/1471-2407-13-240.

41. Raphael MJ, Biagi JJ, Kong W, Mates M, Booth CM, Mackillop WJ. The relationship between time to initiation of adjuvant chemotherapy and survival in breast cancer: a systematic review and meta-analysis. Breast Cancer Res Treat. 2016;160(1):17–28 10.1007/s10549-016-3960-3.

42. Zhan QH, Fu JQ, Fu FM, Zhang J, Wang C. Survival and time to initiation of adjuvant chemotherapy among breast cancer patients: a systematic review and meta-analysis. Oncotarget. 2018;9(2):2739–51 10.18632/oncotarget.23086.

43. Des Guetz G, Nicolas P, Perret GY, Morere JF, Uzzan B. Does delaying adjuvant chemotherapy after curative surgery for colorectal cancer impair survival? A meta-analysis. Eur J Cancer. 2010;46(6):1049–55 10.1016/j.ejca.2010.01.020.

44. Biagi JJ, Raphael MJ, Mackillop WJ, Kong W, King WD, Booth CM. Association between time to initiation of adjuvant chemotherapy and survival in colorectal cancer: a systematic review and meta-analysis. JAMA. 2011;305(22):2335–42 10.1001/jama.2011.749.

45. Usón PLS, Bugano DDG, França MS, Antunes YPPV, Taranto P, Kaliks RA, Del Giglio A. Does Time-to-Chemotherapy Impact the Outcomes of Resected Ovarian Cancer? Meta-analysis of Randomized and Observational Data. Int J Gynecol Cancer. 2017;27(2):274–80 10.1097/igc.0000000000000923.

46. Liu Y, Zhang T, Wu Q, Jiao Y, Gong T, Ma X, Li D. Relationship between initiation time of adjuvant chemotherapy and survival in ovarian cancer patients: a dose-response meta-analysis of cohort studies. Sci Rep. 2017;7(1):9461 10.1038/s41598-017-10197-1.

47. Hershman DL, Wang X, McBride R, Jacobson JS, Grann VR, Neugut AI. Delay of adjuvant chemotherapy initiation following breast cancer surgery among elderly women. Breast Cancer Res Treat. 2006;99(3):313–21 10.1007/s10549-006-9206-z.

48. Francois Y, Nemoz CJ, Baulieux J, Vignal J, Grandjean JP, Partensky C, Souquet JC, Adeleine P, Gerard JP. Influence of the interval between preoperative radiation therapy and surgery on downstaging and on the rate of sphincter-sparing surgery for rectal cancer: the Lyon R90-01 randomized trial. J Clin Oncol. 1999;17(8):2396 10.1200/jco.1999.17.8.2396.

49. Berglund A, Cedermark B, Glimelius B. Is it deleterious to delay the start of adjuvant chemotherapy in colon cancer stage III? Ann Oncol. 2008;19(2):400–2 10.1093/annonc/mdm582.

50. André T, Quinaux E, Louvet C, Colin P, Gamelin E, Bouche O, Achille E, Piedbois P, Tubiana-Mathieu N, Boutan-Laroze A, Flesch M, Lledo G, Raoul Y, Debrix I, Buyse M, de Gramont A. Phase III Study Comparing a Semimonthly With a Monthly Regimen of Fluorouracil and Leucovorin As Adjuvant Treatment for Stage II and III Colon Cancer Patients: Final Results of GERCOR C96.1. J Clin Oncol. 2007;25(24):3732–8 10.1200/jco.2007.12.2234.

51. Gray R, Barnwell J, McConkey C, Hills RK, Williams NS, Kerr DJ. Adjuvant chemotherapy versus observation in patients with colorectal cancer: a randomised study. Lancet. 2007;370(9604):2020–9 10.1016/s0140-6736(07)61866-2.

52. Taal BG, Van Tinteren H, Zoetmulder FA. Adjuvant 5FU plus levamisole in colonic or rectal cancer: improved survival in stage II and III. Br J Cancer. 2001;85(10):1437–43 10.1054/bjoc.2001.2117.

53. Cheung WY, Neville BA, Earle CC. Etiology of delays in the initiation of adjuvant chemotherapy and their impact on outcomes for Stage II and III rectal cancer. Dis Colon Rectum. 2009;52(6):1054–63; discussion 64 10.1007/DCR.0b013e3181a51173.

54. Hershman D, Hall MJ, Wang X, Jacobson JS, McBride R, Grann VR, Neugut AI. Timing of adjuvant chemotherapy initiation after surgery for stage III colon cancer. Cancer. 2006;107(11):2581–8 10.1002/cncr.22316.

55. Pronzato P, Campora E, Amoroso D, Bertelli G, Botto F, Conte PF, Sertoli MR, Rosso R. Impact of administration-related factors on outcome of adjuvant chemotherapy for primary breast cancer. Am J Clin Oncol. 1989;12(6):481–5 10.1097/00000421-198912000-00004.

56. Zaheed M, Wilcken N, Willson ML, O’Connell DL, Goodwin A. Sequencing of anthracyclines and taxanes in neoadjuvant and adjuvant therapy for early breast cancer. Cochrane Database of Syst Rev. 2019(2) 10.1002/14651858.CD012873.pub2.

57. Weller D, Vedsted P, Rubin G, Walter FM, Emery J, Scott S, Campbell C, Andersen RS, Hamilton W, Olesen F, Rose P, Nafees S, van Rijswijk E, Hiom S, Muth C, Beyer M, Neal RD. The Aarhus statement: improving design and reporting of studies on early cancer diagnosis. Br J Cancer. 2012;106(7):1262–7 10.1038/bjc.2012.68.

58. Coxon D, Campbell C, Walter FM, Scott SE, Neal RD, Vedsted P, Emery J, Rubin G, Hamilton W, Weller D. The Aarhus statement on cancer diagnostic research: turning recommendations into new survey instruments. BMC Health Services Research. 2018;18(1):677 10.1186/s12913-018-3476-0.

59. Hanna TP, King WD, Thibodeau S, Jalink M, Paulin GA, Harvey-Jones E, O’Sullivan DE, Booth CM, Sullivan R, Aggarwal A. Mortality due to cancer treatment delay: systematic review and meta-analysis. BMJ. 2020;371:m4087 10.1136/bmj.m4087.

60. Gheorghe A, Maringe C, Spice J, Purushotham A, Chalkidou K, Rachet B, Sullivan R, Aggarwal A. Economic impact of avoidable cancer deaths caused by diagnostic delay during the COVID-19 pandemic: A national population-based modelling study in England, UK. European Journal of Cancer. 2021;152:233–42 10.1016/j.ejca.2021.04.019.

