## Supplementary Tables 1-3 for "The impact of lag time to cancer diagnosis and treatment on clinical outcomes prior to the COVID-19 pandemic: a scoping review of systematic reviews and meta-analyses"

**Supplementary Table 1.** Search strategy used to identify relevant systematic reviews and meta-analyses on the association between time to cancer diagnosis and treatment and outcomes of interest

| Search term | Search number | Keywords/MeSH terms |
|---|---|---|
| Cancer | 1 | 'neoplasm'/exp OR (Neoplasms) OR (Cancer) OR (Cancers) OR (Neoplasia) OR (Neoplasm) OR (Tumors) OR (Tumor) OR (Malignancy) OR (Malignancies) OR (Malignant Neoplasms) OR (Malignant Neoplasm) OR (Neoplasm Benign) |
| Diagnosis & Treatment | 2 | (Diagnosis) OR (diagnose) OR (treat) OR (treatment) OR (therapy) OR (care) OR (screen) OR (surgery) OR (radiation therapy) OR (systemic therapy) OR (chemotherapy) OR (adjuvant chemotherapy) OR (adjuvant radiotherapy) OR (neoadjuvant chemotherapy) OR (neoadjuvant radiotherapy) |
| Exposure | 3 | (delay) OR (wait time) or (postpone) OR (interval) OR (deferred) OR (deferral) OR (time to surgery) OR (time to treatment) OR (waiting period) OR (patient delay) OR (provider delay) OR (doctor delay) OR (time to treatment initiation) OR (system delay) OR (doctor delay) OR (professional delay) OR (time interval) OR (postponement) OR (time to diagnosis) |
| Outcome | 4 | (outcomes) OR (recurrence) OR (survival) OR (mortality) OR (tumour progression) OR (postoperative complications) OR (oncologic outcomes) OR (disease free survival) OR (overall survival) OR (pathological complete response) OR (recurrence free survival) OR (local recurrence) OR (metastasis) OR (progression free survival) |
| Study type | 5 | (systematic review) OR (meta-analysis) OR (metanalyses) OR (systematic reviews) |
| Aggregation of search terms | 6 | 1 AND 2 AND 3 AND 4 AND 5 |
| Limits | 7 | Limit 6 to yr="2010-2019" |

The search was performed on 15 February 2021, limiting to publications from before the COVID-19 pandemic (1 January 2010 - 31 December 2019), with no restriction on publication language.

**Supplementary Table 2.** Characteristics of included systematic reviews on the association between time to cancer diagnosis and treatment and clinical outcomes

| First author (year) | Databases searched | Number of hits | Number of included studies | Total Number of participants | Countries in which studies were conducted |
|---|---|---|---|---|---|
| Brasme (2012) (28) | EMBASE, Science Citation Index, Google Scholar | 6,412 | 98 | 22,619 | Not reported |
| van den Bergh (2013) (24) | PubMed and EMBASE | 4,950 | 17 | 34,517 | Not reported |
| Doubeni (2018) (30) | MEDLINE | 4,979 | 8 | 77,035 | Canada, France, USA |
| Foster (2013) (26) | MEDLINE, EMBASE | 2,111 | 16 | 2,628 | Not reported |
| Lethaby (2013) (29) | Medline, EMBASE, EMBASE Classic, Centre for Reviews and Dissemination databases, Cochrane Library, Medline In-Process, Other Non-Indexed Citations | 1,665 | 32 | 11,016 | France, Denmark, Israel, UK, Japan, Switzerland, Germany, Canada, USA, China, Brazil, Belgium, Italy, Hong Kong, Singapore, Turkey, South Africa, Nigeria, Sweden |
| Neal (2015) (20) | MEDLINE, MEDLINE in-process, EMBASE, Cumulative Index to Nursing and Allied Health Literature, PsychINFO, Cochrane Central Register of Controlled Trials, Database of Abstracts of Reviews of Effects, Cochrane Database of Systematic Reviews, Health Technology Assessment Database, NHS Economic Evaluation database. | 193,077 | 117 | 401,706 | UK, Italy, Spain, Netherlands, Denmark, Finland, France, Norway, Switzerland, Sweden, Germany, Poland, Austria, Belgium, Romania, Greece, Denmark, India, Japan, China, Hong Kong, Malaysia, South Korea, USA, Canada, Brazil, Turkey, Israel, Australia, New Zealand, Saudi Arabia, Libya, South Africa |
| Graboyes (2018) (23) | PubMed, EMBASE, Cumulative Index to Nursing and Allied Health Literature, Cochrane Library | Not reported | 5 | Not reported | Netherlands, USA, Canada, South Korea |
| Hansen (2018) (22) | PubMed, EMBASE, Cumulative Index to Nursing and Allied Health Literature, Cochrane Library | 3,259 | 5 | 13,514 | Denmark, USA, Canada, South Korea |
| Mattosinho (2019) (27) | PubMed/MEDLINE, Literatura Latino-Americana e do Caribe em Ciências da Saúde, Scientific Electronic Library Online | 434 | 9 | 1,560 | Brazil, Honduras, Chile, Argentina, Pero, México |
| Warren (2019) (25) | PubMed, EMBASE, MEDLINE | 575 | 10 | 30,298 | Not reported |

**Supplementary Table 3.** Characteristics of, and subgroup and/or sensitivity analysis reported by included meta-analyses on the association between time to cancer diagnosis and treatment and clinical outcomes

| First author (year) | Databases searched | Number of hits | Number of included studies | Total number of participants | Subgroup and/or Sensitivity analyses | | | Countries in which studies were conducted |
|---|---|---|---|---|---|---|---|---|
| | | | | | **Covariate** | **Response variable of interest** | **Pooled risk estimate [95% CI] (model type, heterogeneity statistics $I^2$ or Ri)** | |
| Gomez (2009) (37) | MEDLINE, EMBASE, and ISI proceedings | Not reported | 9 | 1,895 | Cancer type | TNM (Oral) | **OR:1.47 [1.09–1.99]** (fixed-effects) | USA, Israel, Canada, Finland, Greece, Thailand, Japan, Brazil, UK |
| | | | | | | | **OR:1.55 [0.96–2.51]** (random-effects, Ri = 0.55) | |
| | | | | | | TNM (Oral delay >1 month) | **OR:1.69 [1.26–2.77]** (fixed-effects) | |
| | | | | | | | **OR:1.41 [0.73–2.75]** (random-effects, Ri = 0.80) | |
| | | | | | Study Quality | TNM (High-quality) | **OR: 1.40 [1.11–1.76] (fixed-effects)** | |
| | | | | | | | OR: 1.31 [0.83–2.07] (random-effects, Ri= 0.74) | |
| | | | | | | TNM (low-quality) | OR: 1.01 [0.62–1.65] | |
| | | | | | | | OR: 1.14 [0.48–2.74] (random-effects, Ri= 0.39) | |
| | | | | | Follow-up period | TNM (≥ 10 year) | **OR: 1.40 [1.11–1.76]** | |
| | | | | | | | OR: 1.31 [0.83–2.07] (random-effects, Ri=0.74) | |
| | | | | | | TNM (< 10 year) | OR: 1.01 [0.62–1.65] | |
| | | | | | | | OR: 1.14 [0.48–2.74] (random-effects, Ri= 0.67) | |
| | | | | | Stratification by site | TNM (Yes) | **OR: 1.51 [1.19–1.90]** | |
| | | | | | | | OR: 1.47 [0.93–2.32] (random-effects, Ri= 0.73) | |
| | | | | | | TNM (No) | OR: 0.85 [0.55–1.31] | |
| | | | | | | | OR: 0.93 [0.49–1.79] (random-effects, Ri= 0.54) | |
| | | | | | Confounding adjustment (tobacco smoking, alcohol consumption) | TNM (Yes) | OR: 1.06 [0.83–1.35] | |
| | | | | | | | OR: 1.02 [0.72–1.43] (random-effects, Ri= 0.46) | |
| | | | | | | TNM (No) | OR: 2.26 [1.54–3.32] | |
| | | | | | | | OR: 2.13 [0.97–4.67] (random-effects, Ri= 0.74) | |
| Des Guetz (2010) (43) | PubMed | 42 | 11 | 17,645 | Excluding the 2 largest studies | Mortality | **RR:1.14 [1.04–1.25]** | Not reported |
| | | | | | Cut-off delays between surgery and AC | Mortality | **RR:1.18 [1.13–1.23]** | |
| | | | | | | Mortality (Colon) | **HR: 1.25 [1.17-1.34]** | |

| | | | | | | | | |
|---|---|---|---|---|---|---|---|---|
| | | | | | Studies on Colon cancer | Mortality (Stage III colon) | **HR: 1.25 [1.18-1.33]** | |
| Biagi (2011) (44) | MEDLINE, EMBASE, Cochrane Database of Systematic Reviews, Cochrane Central Register of Controlled Trials | 198 | 10 | 15,410 | Excluding the 2 largest studies | Mortality | **HR: 1.15 [1.10-1.22]** | Not reported |
| | | | | | Excluding the 3 largest studies | Mortality | **HR: 1.13 [1.10-1.17]** | |
| | | | | | Excluding the 3 largest studies | Cancer-specific survival | **HR: 1.15 [1.10-1.19]** | |
| Seoane (2012) (19) | MEDLINE, EMBASE, ISI Proceedings | 1,016 | 10 | 1,286 | Cancer type | Mortality (oral cancer) | RR: 1.00 [0.92-1.10] (fixed-effects) | Finland, Spain, USA, Denmark |
| | | | | | | | RR: 1.27 [0.81–1.98] (random-effects, Ri = 0.94) | |
| | | | | | | Mortality (pharynx cancer) | **RR: 1.68 [1.22-2.31]** (fixed-effects) | |
| | | | | | | | **RR: 1.69 [1.05–2.72]** (random-effects, Ri = 0.55) | |
| | | | | | | Mortality (larynx cancer) | **RR: 1.05 [1.02-1.08]** (fixed-effects) | |
| | | | | | | | RR: 1.64 [0.91–2.96] (random-effects, Ri = 1.00) | |
| | | | | | Percentage of cases in stage III and IV | Mortality (≥ 60%) | **RR: 1.74 [1.30-2.33]** (fixed-effects) | |
| | | | | | | | **RR: 1.76 [1.21–2.54]** (random-effects, Ri = 0.37) | |
| | | | | | | Mortality (<60%) | **RR: 1.04 [1.01-1.07]** (fixed-effects) | |
| | | | | | | | RR: 1.19 [0.99–1.44] (random-effects, Ri = 0.96) | |
| | | | | | Study design | Mortality (retrospective) | **RR: 1.09 [1.00-1.19]** (fixed-effects) | |
| | | | | | | | **RR: 1.57 [1.11–2.24]** (random-effects, Ri = 0.92) | |
| | | | | | | Mortality (partially prospective) | **RR: 1.04 [1.01-1.07]** (fixed-effects) | |
| | | | | | | | RR: 1.34 [0.69–2.61] (random-effects, Ri = 1.00) | |
| | | | | | Primary care centres | Mortality | **RR: 1.50 [1.25-1.79]** (fixed-effects) | |
| | | | | | | | **RR: 1.77 [1.14–2.73]** (random-effects, Ri = 0.81) | |
| | | | | | Questionnaires | Mortality | **RR: 1.04 [1.01-1.07]** (fixed-effects) | |
| | | | | | | | RR: 1.04 [0.95–1.13] (random-effects, Ri = 0.84) | |
| | | | | | Study participants | Mortality (population-based) | RR: 1.02 [0.92-1.11] (fixed-effects) | |
| | | | | | | | RR: 1.16 [0.82–1.65] (random-effects, Ri = 0.89) | |
| | | | | | | Mortality (hospital-based) | **RR: 1.05 [1.02-1.08]** (fixed-effects) | |
| | | | | | | | **RR: 1.67 [1.14–2.44]** (random-effects, Ri = 0.99) | |
| | | | | | | Mortality | **RR: 1.09 [1.00-1.19]** (fixed-effects) | |

| | | | | | | | | |
|---|---|---|---|---|---|---|---|---|
| | | | | | Source of mortality data unknown | | **RR: 1.68 [1.13–2.49]** (random-effects, Ri = 0.94) | |
| | | | | | Source of data unknown | Mortality | **RR: 1.04 [1.01-1.07]** (fixed-effects) | |
| | | | | | | | RR: 1.17 [0.85–1.60] (random-effects, Ri = 0.99) | |
| | | | | | Adjusted for Sex | Mortality (yes) | **RR: 1.04 [1.01-1.07]** (fixed-effects) | |
| | | | | | | | RR: 1.16 [0.99–1.36] (random-effects, Ri = 0.94) | |
| | | | | | | Mortality (no) | **RR: 2.77 [1.81-4.24]** (fixed-effects) | |
| | | | | | | | **RR: 2.77 [1.81–4.24]** (random-effects, Ri = 0.00) | |
| | | | | | Study quality | Mortality (study quality score ≥4) | **RR: 1.54 [1.28-1.86]** (fixed-effects) | |
| | | | | | | | **RR: 1.77 [1.14–2.75]** (random-effects, Ri = 0.81) | |
| | | | | | | Mortality (study quality score <4) | **RR: 1.04 [1.012-1.07] (**fixed-effects) | |
| | | | | | | | RR: 1.04 [0.93–1.17] (random-effects, Ri = 0.89) | |
| Yu (2013) (40) | PubMed, EMBASE, Cochrane Library, Web of Science | 1,157 | 7 | 34,097 | Stepwise exclusion | Mortality (1 study) | **HR: 1.17 [1.12-1.22]** | Italy, France, Denmark, USA |
| | | | | | | Mortality (2 studies) | **HR: 1.23 [1.12-1.34]** | |
| Seoane (2015) (18) | MEDLINE, EMBASE, Web of Science | 653 | 10 | 1,023 | Study quality | TNM, T1, T2, T4, or T11, high quality | **RR: 2.44 [1.36–4.36]** (fixed-effects) | Canada, Thailand, Finland, Netherlands, UK, Argentina, Spain |
| | | | | | | | **RR: 2.44 [1.36–4.36]** (random-effects, Ri = 0.00) | |
| | | | | | | TNM, T1, T2, T4, or T11, low quality | **RR: 1.53 [1.26–1.86]** (fixed-effects) | |
| | | | | | | | **RR: 1.53 [1.12–2.08]** (random-effects, Ri = 0.55) | |
| Wang (2015) (33) | PubMed, Cochrane Library, EMBASE | 3,053 | 15 | 4,431 | Time interval | pCR rate (5 weeks) | RR: 0.67 [0.40-1.12] | Not reported |
| | | | | | | pCR rate (6 weeks) | RR: 1.03 [0.76-1.42] | |
| | | | | | | pCR rate (7 weeks) | **RR: 1.45 [1.18-1.78]** | |
| | | | | | | pCR rate (8 weeks) | **RR: 1.49 [1.15-1.92]** | |
| | | | | | | pCR rate (10 weeks) | RR: 0.83 [0.65-1.06] | |
| | | | | | | pCR rate (12 weeks) | RR: 0.81 [0.60-1.08] | |
| Gupta (2016) (31) | MEDLINE, EMBASE, Cochrane Register of Controlled Trials | Not reported | 11 included in primary analysis (34 included in review) | 79,616 (of the 34 studies) | Treatment sequencing vs non-sequencing studies | LR (sequencing) | RR: 1.08 [0.98-1.19] (fixed-effects) | Not reported |
| | | | | | | LR (non-sequencing) | **RR: 1.08 [1.01-1.15] (fixed-effects)** | |
| | | | | | | Mortality (sequencing) | RR: 1.0 [0.94-1.06] (fixed-effects) | |
| | | | | | | Mortality (non-sequencing) | RR: 0.98 [0.87-1.10] (fixed-effects) | |
| | | | | | | LR | **RR: 1.07 [1.03-1.10]** | |

| | | | | | | | | |
|---|---|---|---|---|---|---|---|---|
| | | | | | Inclusion of all studies regardless of quality | Mortality | **RR: 1.06 [1.04-1.07]** | |
| Petrelli (2016) (34) | PubMed, EMBASE, Web of Science, Cochrane Library | 5,065 | 13 | 3,584 | -- | | | Not reported |
| Raphael (2016) (41) | MEDLINE, EMBASE | 1,326 | 14 | 56,269 | Study type | Mortality (RCT) | RR: 1.06 [0.97-1.16] (random effects) | Denmark, Italy, France, Canada, US, England, New Zealand |
| | | | | | | Mortality (institution-based cohort study) | **RR: 1.20 [1.00-1.44]** (random effects) | |
| | | | | | | Mortality (population-based cohort study) | **RR: 1.43 [1.43-1.69]** (random effects) | |
| | | | | | Validity | Worsened DFS (high validity only) | **RR: 1.04 [1.00-1.08]** | |
| | | | | | | Worsened DFS (not high validity) | RR: 1.06 [0.97-1.15] | |
| | | | | | | Worsened DFS (high validity) | RR: 1.06 [0.99 -1.12] (random effects) | |
| | | | | | | Worsened DFS (not high validity) | RR: 1.06 [0.97 -1.15] (random effects) | |
| Du (2017) (36) | PubMed, EMBASE, and Cochrane Library | 1,914 | 13 | 19,652 | Stepwise exclusion | pCR | **RR: 1.25 [1.16-1.35]** | Not reported |
| | | | | | | | **RR: 1.25 [1.16-1.35]** | |
| | | | | | | | **RR: 1.31 [1.07-1.60]** | |
| | | | | | | | **RR: 1.25 [1.16-1.35]** | |
| | | | | | | | **RR: 1.26 [1.16-1.36]** | |
| | | | | | | | **RR: 1.27 [1.17-1.37]** | |
| | | | | | | | **RR: 1.25 [1.16-1.35]** | |
| | | | | | | | **RR: 1.25 [1.16-1.36]** | |
| | | | | | | | **RR: 1.25 [1.16-1.35]** | |
| | | | | | | | **RR: 1.25 [1.15-1.35]** | |
| | | | | | | | **RR: 1.24 [1.15-1.35]** | |
| | | | | | | | **RR: 1.26 [1.17-1.36]** | |
| | | | | | Waiting interval | pCR (7-weeks) | **RR: 1.51 [1.20-1.89]** | |
| Lin (2017) (38) | MEDLINE, EMBASE, Clinical Trials, Cochrane Central Register of Controlled Trials. | 2,622 | 5 | 1,016 | Stepwise exclusion | 2-year OS, 5-year OS, pCR, postoperative mortality, anastomotic leakage | -- | Italy, USA, Taiwan, France |

| | | | | | | | | |
|---|---|---|---|---|---|---|---|---|
| Liu (2017) (46) | PubMed, Web of Science | 5,277 | 14 | 59,569 | Study design | Mortality (prospective) (Categorical lag-time variable) | **HR: 1.22 [1.02-1.46] (random-effects, $I^2$ = 48.7%)** | (Continent only provided) Asia, North America, Europe |
| | | | | | | Mortality (retrospective) (Categorical lag-time variable) | **HR: 1.08 [1.01-1.15] (random-effects, $I^2$ = 0%)** | |
| | | | | | Geographic location | Mortality (Asia) (Categorical lag-time variable) | HR: 1.06 [0.80-1.38] (random-effects, $I^2$ = NA) | |
| | | | | | | Mortality (North America) (Categorical lag-time variable) | HR: 1.11 [0.94-1.32] (random-effects, $I^2$ = 87.7%) | |
| | | | | | | Mortality (Europe) (Categorical lag-time variable) | **HR: 1.25 [1.05-1.29] (random-effects, $I^2$ = 21.6%)** | |
| | | | | | Number of cases | Mortality (<600) (Categorical lag-time variable) | HR: 1.16 [0.83-1.62] (random-effects, $I^2$ = 60.6%) | |
| | | | | | | Mortality (≥600) (Categorical lag-time variable) | **HR: 1.15 [1.02-1.29] (random-effects, $I^2$ = 77.6%)** | |
| | | | | | Residual disease | Mortality (Yes) (Categorical lag-time variable) | HR: 0.69 [0.30-1.60] (random-effects, $I^2$ = NA) | |
| | | | | | | Mortality (No) (Categorical lag-time variable) | HR: 1.35 [0.51-3.57] (random-effects, $I^2$ = NA) | |
| | | | | | Chemotherapy | Mortality (Platinum-based) (Categorical lag-time variable) | HR: 1.26 [0.98-1.63] (random-effects, $I^2$ = 7.8%) | |
| | | | | | | Mortality (Platinum-based plus Taxane) (Categorical lag-time variable) | HR: 1.17 [1.04-1.32] (random-effects, $I^2$ = 65.9%) | |
| | | | | | FIGO stage | Mortality (All) (Categorical lag-time variable) | HR: 1.09 [0.93-1.28] (random-effects, $I^2$ = 0%) | |
| | | | | | | Mortality (III–IV) (Categorical lag-time variable) | **HR: 1.23 [1.07-1.42] (random-effects, $I^2$ = 81.5%)** | |

| | | | | | | | |
|---|---|---|---|---|---|---|---|
| | | | | | | Mortality (I–II) (Categorical lag-time variable) | HR: 0.78 [0.51-1.19] (random-effects, $I^2$ = NA) |
| | | | | | Adjustment for FIGO staging as potential confounder | Mortality (Yes) (Categorical lag-time variable) | **HR: 1.16 [1.04-1.30] (random-effects, $I^2$ = 70.9%)** |
| | | | | | | Mortality (No) (Categorical lag-time variable) | HR: 0.92 [0.48-1.77] (random-effects, $I^2$ = NA) |
| | | | | | Adjustment for histology as a potential confounder | Mortality (Yes) (Categorical lag-time variable) | HR: 1.07 [0.91-1.26] (random-effects, $I^2$ = 36.7%) |
| | | | | | | Mortality (No) (Categorical lag-time variable) | **HR: 1.26 [1.19-1.34] (random-effects, $I^2$ = 0%)** |
| | | | | | Adjustment for residual disease as a potential confounder | Mortality (Yes) (Categorical lag-time variable) | **HR: 1.22 [1.10-1.36] (random-effects, $I^2$ = 23.5%)** |
| | | | | | | Mortality (No) (Categorical lag-time variable) | HR: 1.09 [0.89-1.34] (random-effects, $I^2$ = 62.6%) |
| | | | | | Study design | Mortality (prospective) (Continuous lag-time variable) | **HR: 1.06 [1.00-1.12] (random-effects, $I^2$ = 0%)** |
| | | | | | | Mortality (retrospective) (Continuous lag-time variable) | HR: 1.04 [0.97-1.13] (random-effects, $I^2$ = 69.1%) |
| | | | | | Geographic location | Mortality (Asia) (Continuous lag-time variable) | **HR: 1.15 [1.04-1.27] (random-effects, $I^2$ = NA)** |
| | | | | | | Mortality (North America) (Continuous lag-time variable) | **HR: 1.01 [1.00-1.02] (random-effects, $I^2$ = 0%)** |
| | | | | | | Mortality (Europe) (Continuous lag-time variable) | **HR: 1.06 [1.00-1.12] (random-effects, $I^2$ = 0%)** |
| | | | | | Number of cases | Mortality (<600) | **HR: 1.07 [1.00-1.15] (random-effects, $I^2$ = 44.2%)** |

| | | | | | | | |
|---|---|---|---|---|---|---|---|
| | | | | | | (Continuous lag-time variable) | |
| | | | | | | Mortality (≥600) (Continuous lag-time variable) | **HR: 1.01 [1.0-1.02] (random-effects, $I^2$ = 55.8%)** |
| | | | | | Residual disease | Mortality (Yes) (Continuous lag-time variable) | **HR: 1.09 [1.01-1.78] (random-effects, $I^2$ = 0%)** |
| | | | | | | Mortality (No) (Continuous lag-time variable) | HR: 0.98 [0.94-1.03] (random-effects, $I^2$ = 0%) |
| | | | | | Chemotherapy | Mortality (Platinum-based) (Continuous lag-time variable) | HR: 1.00 [0.90-1.11] (random-effects, $I^2$ = NA) |
| | | | | | | Mortality (Platinum-based plus Taxane) (Continuous lag-time variable) | HR: 1.02 [0.99-1.05] (random-effects, $I^2$ = 26.2%) |
| | | | | | | Mortality (NA) (Continuous lag-time variable) | **HR: 1.15 [1.04-1.27] (random-effects, $I^2$ = NA)** |
| | | | | | FIGO stage | Mortality (All) (Continuous lag-time variable) | HR: 1.03 [0.93-1.14] (random-effects, $I^2$ = NA) |
| | | | | | | Mortality (III–IV) (Continuous lag-time variable) | HR: 1.05 [0.99-1.11] (random-effects, $I^2$ = 66.4%) |
| | | | | | Adjustment for FIGO staging as potential confounder | Mortality (Yes) (Continuous lag-time variable) | HR: 1.05 [0.99-1.11] (random-effects, $I^2$ = 66.4%) |
| | | | | | | Mortality (No) (Continuous lag-time variable) | HR: 1.03 [0.93-1.14] (random-effects, $I^2$ = NA) |
| | | | | | Adjustment for histology as a potential confounder | Mortality (Yes) (Continuous lag-time variable) | HR: 1.02 [0.99-1.05] (random-effects, $I^2$ = 26.2%) |
| | | | | | | Mortality (No) (Continuous lag-time variable) | HR: 1.07 [0.94-1.23] (random-effects, $I^2$ = 72.0%) |
| | | | | | Adjustment for residual | Mortality (Yes) | **HR: 1.07 [1.00-1.15] (random-effects, $I^2$ = NA)** |

| | | | | | | | | |
|---|---|---|---|---|---|---|---|---|
| | | | | | disease as a potential confounder | (Continuous lag-time variable) | | |
| | | | | | | Mortality (No) (Continuous lag-time variable) | HR: 1.04 [0.98-1.09] (random-effects, $I^2$ = 54.6%) | |
| Loureiro (2017) (39) | PubMed/MEDLINE, EMBASE, Thomson Reuters, Web of Science Core Collection, ASTRO and ESTRO proceedings of annual meetings (2000-2013) | 1,369 | 12 | 5,212 | Excluding studies without WT to RT period restrictions | Mortality | HR: 1.0 [0.90-1.12] | Spain, Canada, Brazil, USA, Germany |
| Usón (2017) (45) | PubMed | 239 | 12 | 12,056 | Stepwise exclusion | Mortality, 3-year | **OR: 1.17 [1.05-1.29] (fixed-effects)** | Not reported |
| Zhan (2017) (42) | PubMed, EMBASE, Web of Science, Cochrane library, ASCO meeting abstracts | 2,390 | 12 | 78,462 | Excluding the largest 2 studies | Mortality | **HR: 1.10 [1.08-1.12] (random-effects, $I^2$ = 43.5%)** | Italy, France, Denmark, USA, UK, China |
| | | | | | Publication year | Mortality (1999) | **HR: 2.15 [1.2-3.85] (random-effects, $I^2$ = %NA)** | |
| | | | | | | Mortality (2005) | HR: 0.00 [0.02-1.05] (random-effects, $I^2$ = 0%) | |
| | | | | | | Mortality (2006) | **HR: 1.22 [1.11-1.34] (random-effects, $I^2$ = 0%)** | |
| | | | | | | Mortality (2013) | **HR: 1.08 [1.06-1.10] (random-effects, $I^2$ = NA)** | |
| | | | | | | Mortality (2014) | **HR: 1.10 [1.06-1.15] (random-effects, $I^2$ = 0%)** | |
| | | | | | | Mortality (2015) | **HR: 1.32 [1.23-1.41] (random-effects, $I^2$ = 0%)** | |
| | | | | | | Mortality (2016) | **HR: 1.13 [1.08-1.19] (random-effects, $I^2$ = 0%)** | |
| | | | | | Excluding the largest study | Worsened DFS | **HR: 1.09 [1.03-1.14]** | |
| Wu (2018) (32) | PubMed, EMBASE, MEDLINE, Cochrane Library | 897 | 5 | 1,244 | -- | | | Not reported |
| Petrelli (2019) (35) | MEDLINE, EMBASE, Cochrane Library | 8,752 | 34 | 141,853 | -- | | | Canada, US, Sweden, Netherlands, UK, Brazil, Korea, Denmark, Taiwan, Italy, Japan, China |

| | | | | | | | | |
|---|---|---|---|---|---|---|---|---|
| Zhao (2019) (21) | PubMed, EMBASE, Medline, Cochrane, ClinicalTrials.gov | 258 | 8 | 885 | Treatment type | Disease progression (Melphalan-prednisone) | **RR: 0.22 [0.08-0.64] (random-effects, $I^2$ = 59%)** | Not reported |
| | | | | | | Disease progression (bisphosphonate) | RR: 1.00 [0.83-1.21] (random-effects, $I^2$ = 0%) | |
| | | | | | | Disease progression (immunomodulatory drug | **RR: 0.43 [0.31-0.59] (random-effects, $I^2$ = 0%)** | |
| | | | | | | Disease progression (Monoclonal antibody) | RR: 0.87 [0.36-2.07] (random-effects, $I^2$ = NA) | |
| | | | | | Treatment type | Mortality (Melphalan-prednisone) | RR: 1.05 [0.82-1.35] (fixed-effects, $I^2$ = 37%) | |
| | | | | | | Mortality (bisphosphonate) | RR: 0.76 [0.18-3.29] (fixed-effects, $I^2$ = NA) | |
| | | | | | | Mortality (immunomodulatory drug) | RR: 0.63 [0.38-1.04] (fixed-effects, $I^2$ = 34%) | |
| | | | | | | Mortality (Monoclonal antibody) | RR: 0.73 [0.17-3.08] (fixed-effects, $I^2$ = 33%) | |
| | | | | | High-risk SMM | Disease progression | **RR: 0.51[0.37-0.70] (fixed-effects, $I^2$ = 47%)** | |
| | | | | | High-risk SMM | Mortality | **RR: 0.53 [0.29-0.97] (fixed-effects, $I^2$ = 0%)** | |

AC, adjuvant chemotherapy; CI, confidence interval; DFS, disease-free survival; HR, hazard ratio; LR, local recurrence; OR, odds ratio; OS, overall survival; pCR, pathological complete response; RCT, randomized controlled trial; RR, risk ratio; SMM, smoldering multiple myeloma

Significant pooled risk estimates are bolded.
--, indicate that subgroup and/or sensitivity analyses were not conducted or were not available.
